# Randomized Trial of Ketamine Masked by Surgical Anesthesia in Depressed Patients

**DOI:** 10.1101/2023.04.28.23289210

**Authors:** Theresa R. Lii, Ashleigh E. Smith, Josephine R. Flohr, Robin L. Okada, Cynthia A. Nyongesa, Lisa J. Cianfichi, Laura M. Hack, Alan F. Schatzberg, Boris D. Heifets

**Author notes:** Dr. Heifets can be contacted at.

## Abstract

**BACKGROUND:** Ketamine may have antidepressant properties, but its acute psychoactive effects complicate successful masking in placebo-controlled trials.

**METHODS:** In a triple-masked, randomized, placebo-controlled trial, 40 adult patients with major depressive disorder were randomized to a single infusion of ketamine (0.5 mg/kg) or placebo (saline) during anesthesia as usual for routine surgery. The primary outcome was depression severity measured by the Montgomery-Åsberg Depression Rating Scale (MADRS) at 1, 2, and 3 days post-infusion. The secondary outcome was the proportion of participants with clinical response (≥50% reduction in MADRS scores) at 1, 2, and 3 days post-infusion. After all follow-up visits, participants were asked to guess which intervention they received.

**RESULTS:** Mean MADRS scores did not differ between groups at screening or pre-infusion baseline. The mixed-effects model showed no evidence of effect of group assignment on post-infusion MADRS scores at 1 to 3 days post-infusion (−5.82, 95% CI −13.3 to 1.64, p=0.13). Clinical response rates were similar between groups (60% versus 50% on day 1) and comparable to previous studies of ketamine in depressed populations. Secondary and exploratory outcomes did not find statistical separation of ketamine from placebo. 36.8% of participants guessed their treatment assignment correctly; both groups allocated their guesses in similar proportions. One serious adverse event occurred in each group, unrelated to ketamine administration.

**CONCLUSION:** In adults with major depressive disorder, a single dose of intravenous ketamine delivered during surgical anesthesia had no greater effect than placebo in acutely reducing the severity of depressive symptoms. This trial successfully masked treatment allocation in moderate-to-severely depressed patients using surgical anesthesia. While it is impractical to use surgical anesthesia for most placebo-controlled trials, future studies of novel antidepressants with acute psychoactive effects should make efforts to fully mask treatment assignment in order to minimize subject-expectancy bias. (ClinicalTrials.gov number, NCT03861988)

## INTRODUCTION

Ketamine, a dissociative anesthetic with multiple molecular targets^1,2^, is associated with rapid-acting antidepressant effects in patients with major depressive disorder (MDD), including those with treatment-resistant depression (TRD)^3–5^. Across studies, an intravenous infusion of 0.5 mg/kg ketamine produces a clinical response in 41% and remission in 19% of patients with TRD at 24 hours^6^. Therapeutic effects appear within 2 hours of a single ketamine infusion^5^.

In most randomized controlled trials (RCTs) of ketamine for depression, participant masking has been nearly impossible given the drug’s obvious acute psychological effects. Inadequate masking presents a major confound to interpreting studies of ketamine, as well as other rapid-acting psychoactive therapeutics such as psilocybin and methylenedioxymethamphetamine (MDMA)^7–10^, to the extent that most investigations do not report on the success of participant masking^10^. Incomplete masking may lead to subject-expectancy bias, which occurs when a research subject has an expectation for a given result that influences the reported outcome. Subject-expectancy bias may contribute to overestimation of treatment effect sizes in antidepressant trials involving ketamine^11^.

We utilized ketamine’s established safety in surgical settings by conducting a randomized placebo-controlled trial in which the administration of ketamine was masked by other surgical anesthetics. The primary aim of this study was to determine whether ketamine, given at a dose of 0.5 mg/kg over 40 minutes during surgical anesthesia, produces a greater antidepressant effect than placebo. We recruited patients with depression severity comparable to previous studies and analyzed similar follow-up time points. We hypothesized that ketamine is superior to an inert placebo (0.9% sodium chloride, *i.e.,* normal saline) in reducing depression symptoms within the first 3 days post-infusion in a population of adults with moderate-to-severe levels of MDD. A secondary aim was to test whether a conscious dissociative reaction to ketamine is needed for an antidepressant response.

## Methods

### TRIAL OVERSIGHT

This was an investigator-initiated sponsored by the university and the Society for Neuroscience in Anesthesiology and Critical Care. The trial protocol was approved by the institutional review board at Stanford University (Protocol #49114) and all participants gave written informed consent. In this study, a Data Safety Monitoring Board was not required as the primary study intervention did not deviate from standard of care and posed no known incremental risk to participants. Randomization and drug compounding were handled by Stanford Health Care Investigational Drug Service. The first and last authors designed the trial. The first author analyzed the data and wrote the first draft of the manuscript. The second through sixth performed the trial and collected the data. The penultimate author provided content expertise and advised trial design. The overall trial was overseen by the last author. The authors vouch for the accuracy and completeness of the data and for the fidelity of the trial to the protocol. There was no industry involvement in the collection or analysis of the data, and no agreements were in place between the authors and any commercial entity.

### PARTICIPANTS

Adults undergoing elective non-cardiac, non-intracranial surgery were recruited from preoperative clinics at Stanford University Medical Center. The 8-item Patient Health Questionnaire (PHQ-8) was distributed to patients through a perioperative mental health screening service. To be eligible for the study, patients must score > 12 on the PHQ-8, corresponding with at least moderate depression^12^. Research staff screened electronic health records (EHR) of patients scheduled for surgery who scored > 12 points on the PHQ-8; those without exclusion criteria documented in the EHR were introduced to the study and consented for an additional screening visit. Surgical clinics also referred patients with symptomatic depression who expressed interest in the trial. These patients were contacted by research staff for a telephone pre-screen, and qualifying patients were consented for an additional screening visit. At this visit, research staff collected information on demographics, medical, and psychiatric history, including level of antidepressant treatment resistance assessed by the Maudsley Staging Method (MSM)^13^. Inclusion and exclusion criteria were assessed via a hybrid approach of corroborating EHR data with patient self-report.

Inclusion criteria included English literacy, body mass index of 17-40 kg/m^2^, a diagnosis of MDD (single or recurrent) and a major depressive episode of ≥ 4 weeks duration prior to screening. The diagnosis of MDD was confirmed by the Mini International Neuropsychiatric Interview Module A^14^. Participants must also have had a combined score of ≥ 31 from the Montgomery-Åsberg Depression Rating Scale^15^ (MADRS) and the Hospital Anxiety and Depression Scale^16^ (HADS). These scales were administered in-person, or by secure video conference or telephone, which have been validated for the PHQ^17^, HADS^18^, and MADRS^19, 20^.

Exclusion criteria included pregnancy, breastfeeding, moderate or severe substance use disorder, history of schizophrenia or schizoaffective disorder, dementia or other amnestic cognitive disorder, history of surgery involving the brain or meninges, encephalitis, meningitis, or degenerative central nervous system disorder, clinically significant thyroid dysfunction within the past 6 months, and chronic use of ≥ 90 morphine milligram equivalents (MME) per day. Patients at high risk of suicidal behavior on the Columbia-Suicide Severity Rating Scale^21^ were also excluded. Concurrent psychotherapy and antidepressant therapy were allowed if therapy was stable for ≥ 4 weeks prior to screening.

### TRIAL DESIGN AND PROCEDURES

This was a triple-masked, randomized, placebo-controlled trial. Prior to randomization, five patients were recruited for an open label study to evaluate study procedures. Data from these five patients are not included in this manuscript. For the randomized trial, twenty participants were allocated to a single dose of intravenous ketamine (0.5 mg/kg diluted into 40 ml of normal saline, infused over 40 minutes using a programmable pump). Another twenty participants were allocated to 40 ml of normal saline infused similarly over 40 minutes. Pharmacy staff randomized participants using computerized block randomization with 5 blocks of 8. The participant, investigators, and direct care providers (*e.g.,* anesthesiologists) were masked. Unmasking occurred after all 40 participants progressed through all follow-up timepoints (i.e. end of trial).

Processed electroencephalography (EEG) from a SedLine® device (Masimo Corporation, Irvine, California, USA) was used to confirm anesthetic depth, measured by the device’s Patient State Index (PSI). To ensure participant masking, the infusion was initiated after anesthetic induction and surgical incision, during maintenance anesthesia (PSI of 25 to 50, consistent with the manufacturer recommendations for general anesthesia). The study drug was provided to the anesthesiologist in an unlabeled syringe.

Anesthesiologists, masked to patient group allocation, administered routine anesthesia tailored to the surgical procedure and patient comorbidities; they were asked to avoid altering the anesthetic in response to any perceived changes to the processed EEG during the study drug infusion (excepting excessively high PSI values indicating the patient was at risk of intraoperative awareness). Anesthesiologists were asked to minimize use of nitrous oxide (N_2_O), which has reported antidepressant effects^22^. Agents used for anesthetic maintenance included intravenous propofol and inhaled sevoflurane or isoflurane. A standard multimodal analgesic regimen was used, consisting of intravenous opioid and acetaminophen, with or without intravenous lidocaine. Due to the heterogeneity of surgical cases represented in this study, we did not mandate specific anesthetic or analgesic regimens outside of the requested constraint on depth of anesthesia.

### OUTCOME MEASURES

The primary outcome was the MADRS score measured 1, 2, and 3 days post-infusion, as previous studies have found the greatest antidepressant effect occurs within the first 72 hours of a single ketamine infusion^23^. The same sample was assessed at 1, 2 and 3 days post-infusion, as participant dropout only occurred after outcomes were assessed on day 3. Additional assessments were made 5, 7, and 14 days post-infusion and used for exploratory analyses. The MADRS is a clinical rating scale used widely in antidepressant trials; it consists of 10 items which measure the severity of depression in individuals, with a total score ranging from 0 to 60, and higher scores indicating more severe depression^15^. Trained clinical research personnel administered the MADRS.

The secondary outcome was clinical response, defined as ≥50% reduction in MADRS scores from screening baseline^24^. Remission, defined as MADRS score ≤12 in our study^25^, was treated as an exploratory outcome. Other exploratory outcomes included the HADS score, postoperative pain intensity and cumulative opioid use. The HADS is a self-reported questionnaire used to assess the severity of anxiety and depression in hospital patients; it consists of 14 items (7 items measuring anxiety, 7 items measuring depression) with a total score ranging from 0 to 42, with higher scores indicating more severe symptoms^16^. Postoperative pain was assessed by the Brief Pain Inventory Short Form modified for postoperative use (BPI-SF)^26, 27^. The BPI-SF measures severity of pain and its impact on daily functioning; it consists of 9 items, each using a numeric rating scale from 0-10. Inpatient postoperative opioid use, calculated as total daily MME^28^, was abstracted from the EHR for each day of hospitalization. Outpatient postoperative opioid use, *via* pill counts, was obtained during remote assessments. At 14 days post-infusion, participants were asked to guess their treatment arm. Outcomes were assessed in person during postoperative hospitalization, and by video or telephone after discharge.

### STATISTICAL METHODS

Intention-to-treat (ITT) analysis was performed for the primary outcome. A mixed-effects model for repeated measures was the analysis strategy pre-registered before data unmasking to evaluate the antidepressant superiority of ketamine to placebo on postoperative days 1, 2, and 3. The following fixed effects were included in the model: group, time in days, and the interaction between group and time. We included random effects for intercepts and slopes to account for variation in MADRS scores and differential treatment effects. An alternative, non-prespecified mixed-effects model using change from pre-infusion baseline scores on the day of surgery was also used to analyze the primary outcome. An unstructured covariance matrix was used in all mixed models described in this study. We calculated Cohen’s kappa statistic, in a post-hoc analysis, to assess the level of agreement between groups regarding their guesses on treatment allocation. We also performed a simple logistic regression to investigate the relationship between final MADRS score and patients’ treatment group guess (coded as ‘1’ for guessing “ketamine” and ‘0’ otherwise). Both variables were obtained at day 14 during the final assessment. Logistic regression results are reported as odds ratios. All analyses were performed using RStudio software (version 2022.07.1 for MacOS). The lme4 package was used for mixed-effects modeling.

Our sample size estimation was derived from an *a priori* power analysis for the primary outcome. In a randomized controlled trial of ketamine versus active placebo performed by Phillips *et al.*, participants had a mean decrease of 10.9 points (standard deviation [SD] 8.9) in MADRS total score relative to pre-infusion scores compared with a mean decrease of 2.8 points (SD 3.6) with midazolam^29^. For reference, the minimum clinically important difference on the MADRS is estimated to range from 3 to 9 points^30^. Using these results, we computed an estimated total sample size of 38 participants at a two-sided alpha level of 0.05 and 80% power to detect this difference if using parametric testing. An additional 2 participants were added to account for potential attrition, for a total of 40 participants. Interim analyses were not performed.

## Results

### PARTICIPANTS

Participant recruitment occurred between February 2020 and August 2022. The first patient was enrolled in the randomized trial on February 19, 2020, and the final patient was enrolled on August 18, 2022. The last day of follow-up was September 9, 2022. For participant flow through the clinical trial, see the CONSORT diagram (**Figure 1**). The screening visit occurred between 27 days and 16 hours prior to surgery (mean (SD) 5.1 (4.6) days). The mean age of trial participants was 51 years; they were mostly female (70%), white (65%), non-Hispanic (87.5%), employed (62.5%), and never smoked (65%). At screening, both groups had moderate levels of depression (ketamine: mean MADRS score = 27.7, placebo: 30.6) and moderate levels of treatment resistance^13^ (ketamine: mean MSM score = 8.3, placebo: 7.5). The presence of symptomatic depression was also supported by the HADS (ketamine: mean score = 24.6, placebo = 24.7). Although current MDD episode durations were longer in ketamine group, the difference did not reach significance (ketamine: median = 38 months, placebo: 17). Both groups also scored similarly on the BPI-SF at screening, except for two questions in which participants in the ketamine group reported having more pain interference with sleep (mean 7.7 versus 5.5, p=0.02) and enjoyment of life (7.6 versus 5.7, p=0.04). Other characteristics were similar between groups (**Table 1**).

**Figure 1.**
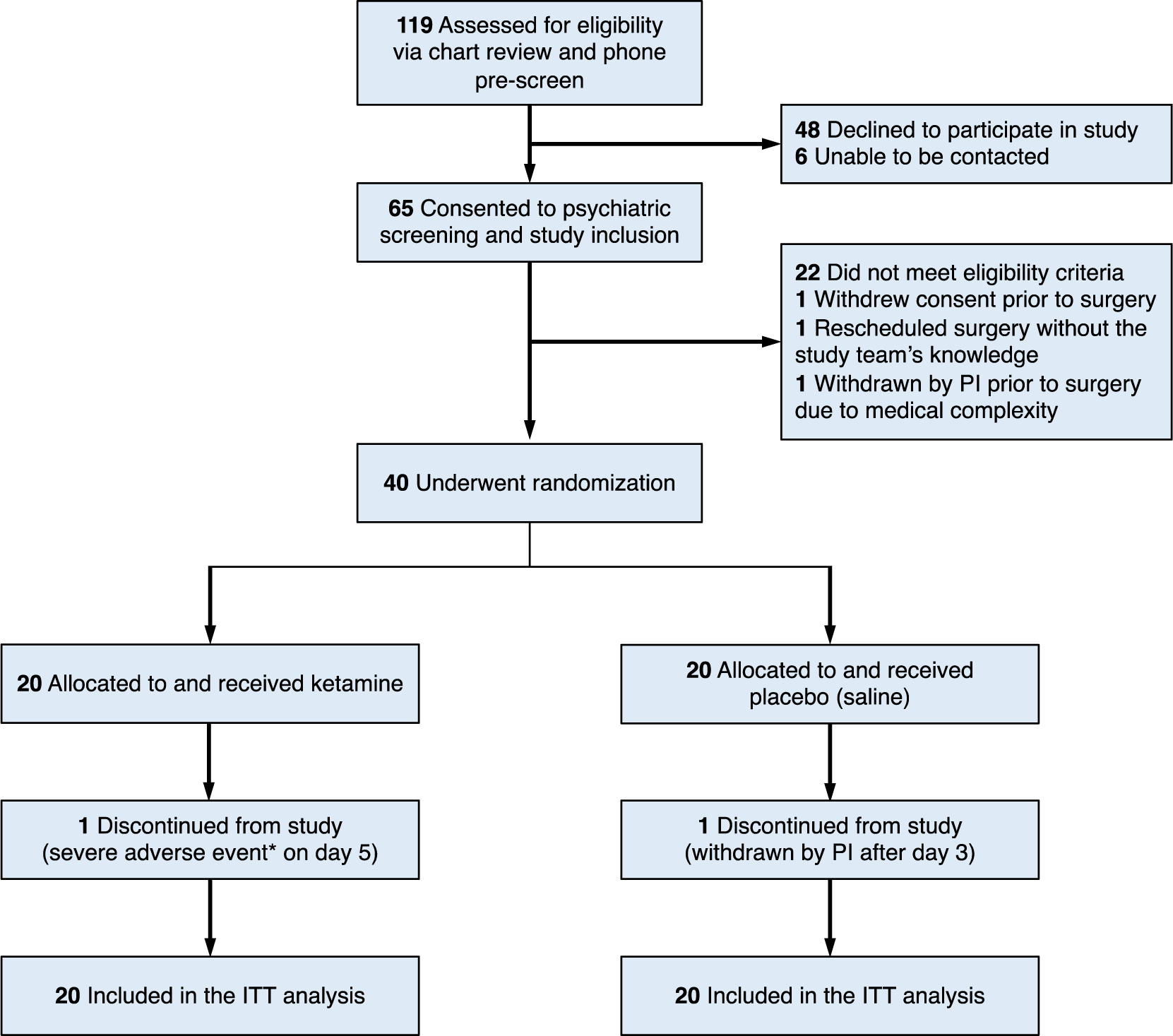
CONSORT Flow Diagram. CONSORT, Consolidated Standards of Reporting Trials. PI, principal investigator. ITT intention-to-treat. Participants were randomized to a single intravenous dose of either ketamine or saline, given during surgical anesthesia. *The severe adverse event refers to an unexpected death that occurred 2 days after the patient was discharged home without complications on postoperative day 3; this patient experienced a witnessed cardiac arrest, attributed to the participant’s medical factors and not resulting from study procedures. †One participant was withdrawn by the PI due to an unanticipated surgical revision of an implanted device, which took place on postoperative day 3 after study assessments were completed.

**Table 1.**
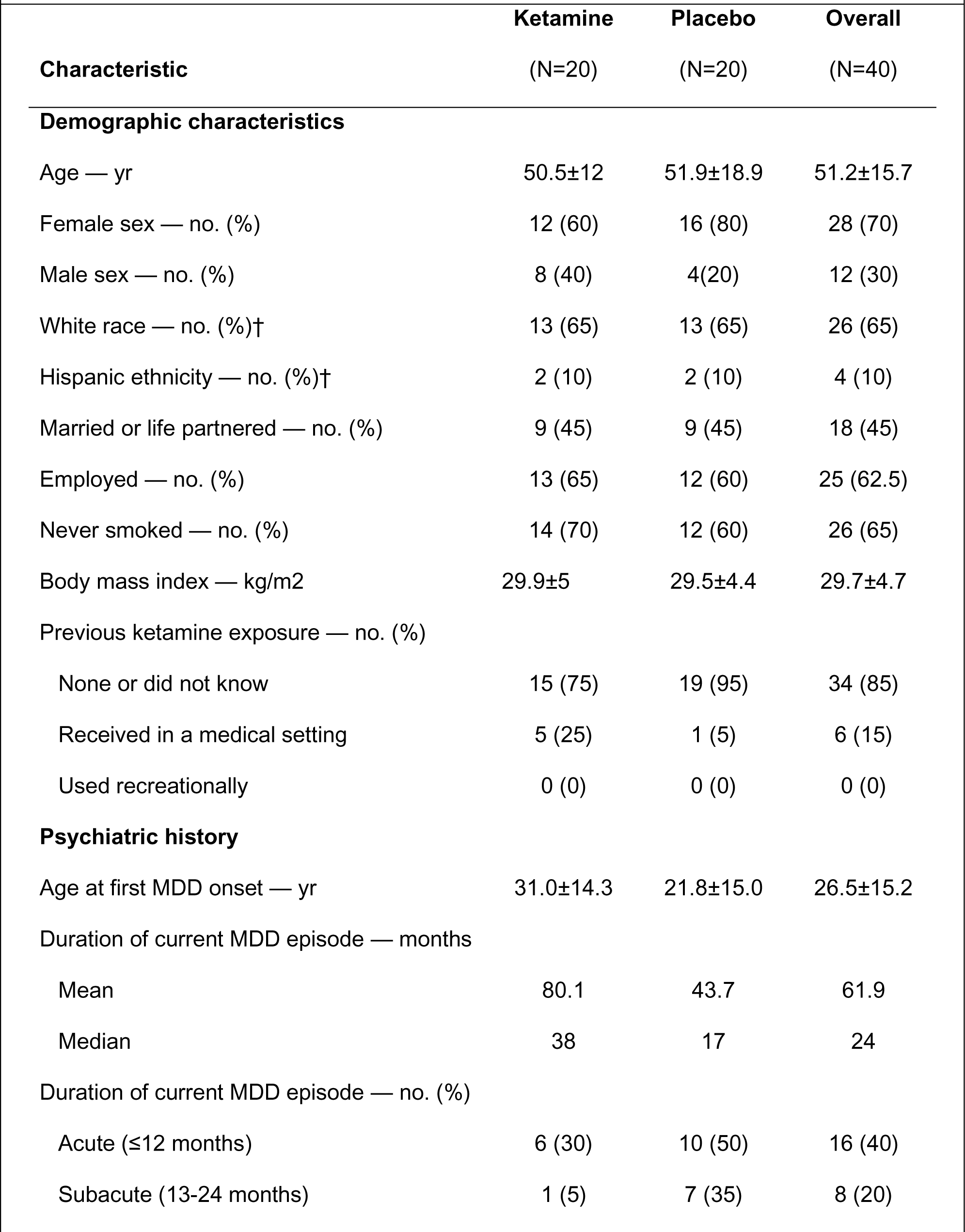
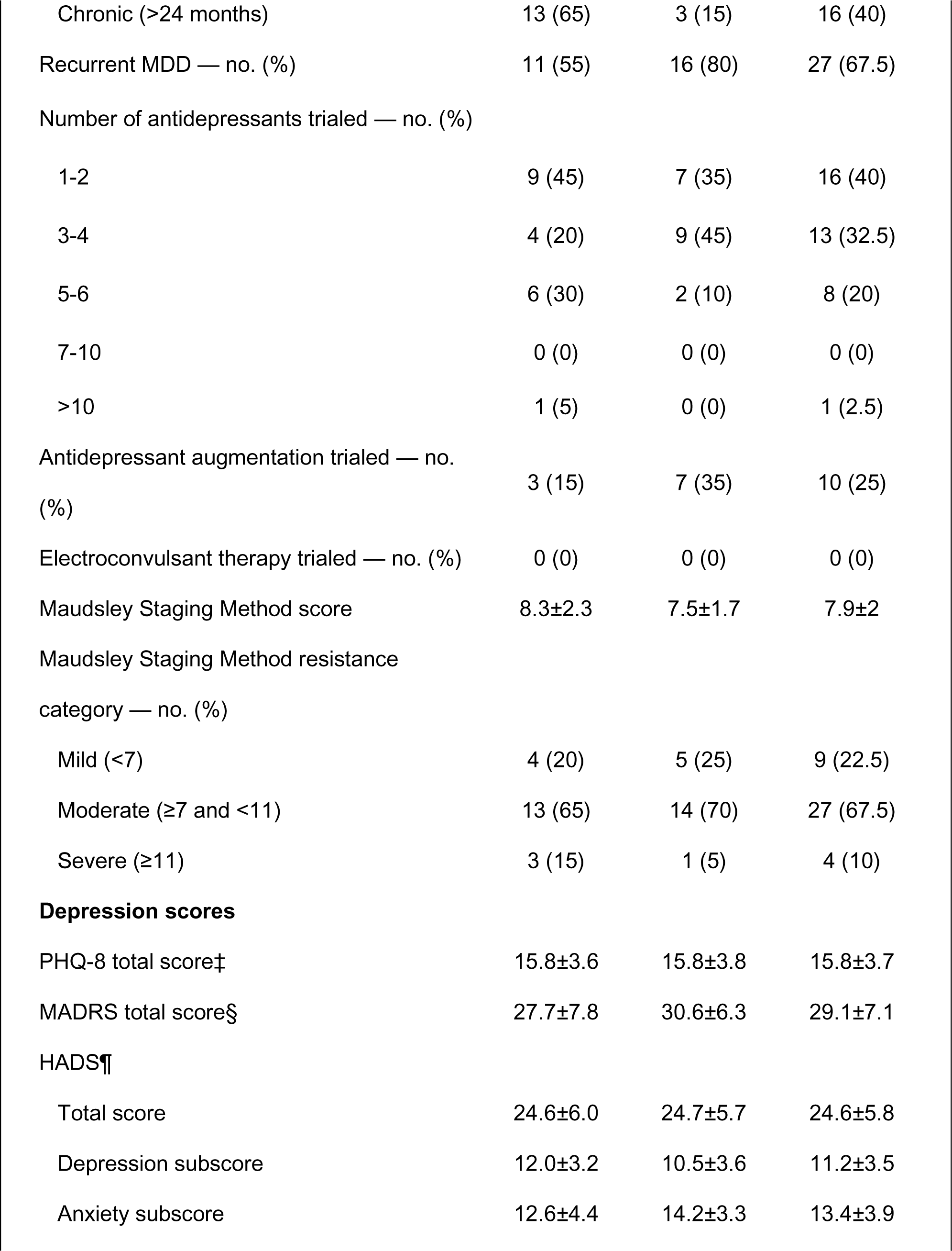
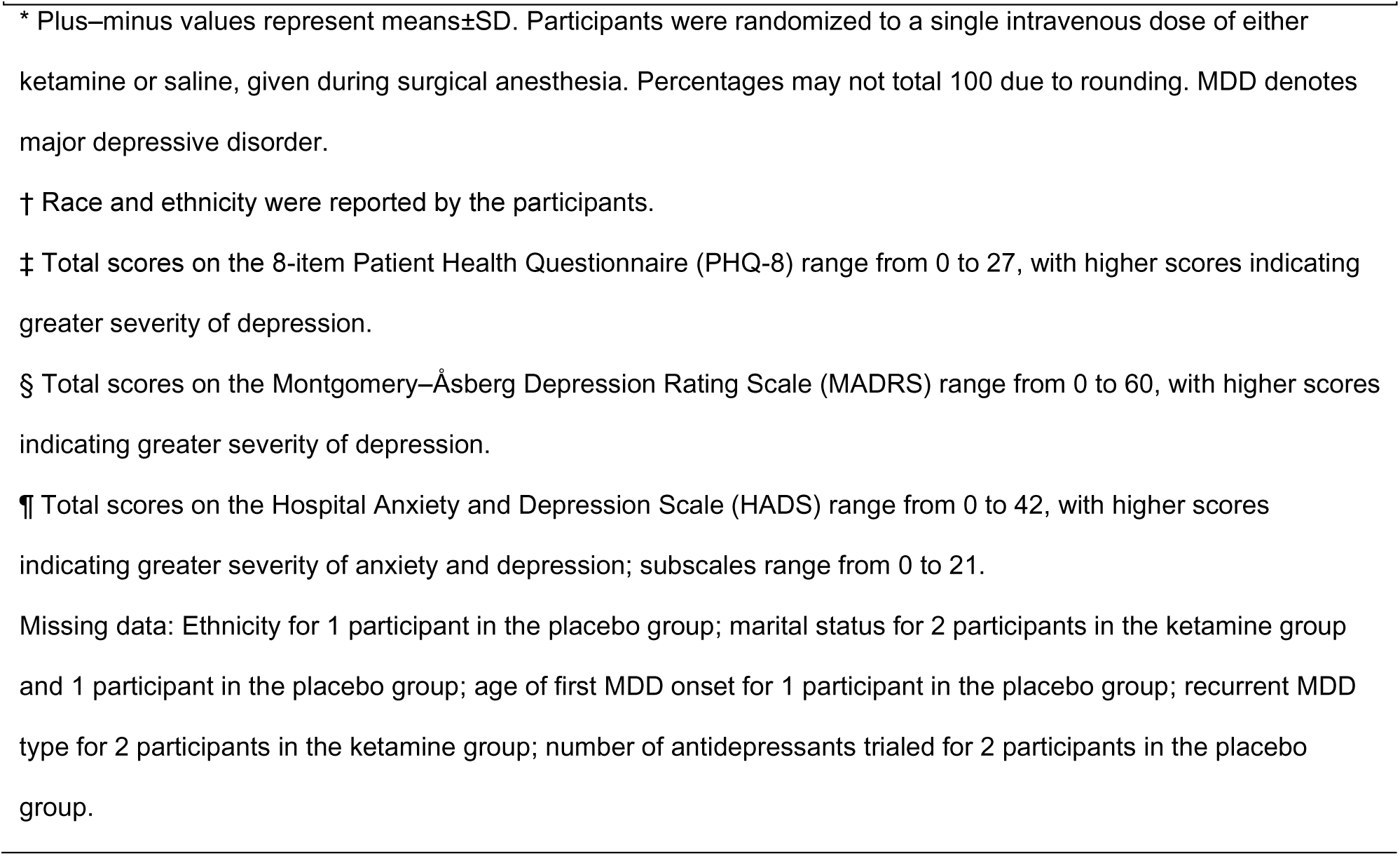
Demographics and Clinical Characteristics of the Participants at Baseline*.

**Table 2** summarizes participants’ surgical and anesthetic characteristics. Participants in both study arms had similar levels of disease burden, as measured by the American Society of Anesthesiologists (ASA) Physical Status Classification^31^, as well as the Charlson Comorbidity Index^32^. Patients presented to a range of surgical departments which were distributed similarly between the two groups. With regards to anesthetic type, all except one underwent general anesthesia. One participant in the ketamine group had monitored anesthesia care with a neuraxial block; however, the depth of anesthesia was within study parameters. Two participants in the ketamine arm and one participant in the placebo arm received N_2_O at a concentration of ≥ 50% for ≥1 hour. Use of preoperative and intraoperative opioids and length of surgery were similar between groups.

**Table 2.** Surgical and Anesthetic Characteristics of the Participants*.

| **Characteristic** | **Ketamine**<br>**(N=20)** | **Placebo**<br>**(N=20)** | **Overall**<br>**(N=40)** |
| --- | --- | --- | --- |
| ASA Physical Status Classification — no. (%)† |  |  |  |
| I | 0 (0) | 1 (5) | 1 (2.5) |
| II | 12 (60) | 15 (75) | 27 (67.5) |
| III | 8 (40) | 4 (20) | 12 (30) |
| CCI total score‡ |  |  |  |
| Mean | 2.1±2.7 | 2.7±2.6 | 2.4±2.7 |
| Median | 1 | 2.5 | 1.5 |
| Surgery department — no. (%) |  |  |  |
| General surgery | 6 (30) | 3 (15) | 9 (22.5) |
| Orthopedics, non-spine | 5 (25) | 4 (20) | 9 (22.5) |
| Otolaryngology | 2 (10) | 2 (10) | 4 (10) |
| Gynecology | 2 (10) | 3 (15) | 5 (12.5) |
| Urology | 2 (10) | 0 (0) | 2 (5) |
| Neurosurgery, non-intracranial | 1 (5) | 3 (15) | 4 (10) |
| Orthopedics, spine | 1 (5) | 2 (10) | 3 (7.5) |
| Thoracic | 1 (5) | 0 (0) | 1 (2.5) |
| Plastics | 0 (0) | 3 (15) | 3 (7.5) |
| Anesthesia type — no. (%) |  |  |  |
| General anesthesia | 19 (95) | 20 (100) | 39 (97.5) |
| Neuraxial block with sedation | 1 (5) | 0 (0) | 1 (2.5) |
| Maintenance anesthetics used — no. (%) |  |  |  |
| Nitrous oxide§ | 2 (10) | 1 (5) | 3 (7.5) |
| Propofol — no. (%) | 18 (90) | 17 (85) | 35 (87.5) |
| Sevoflurane — no. (%) | 17 (85) | 14 (70) | 31 (77.5) |
| Isoflurane — no. (%) | 1 (5) | 3 (15) | 4 (20) |
| Use of regional anesthesia — no. (%) | 1 (5) | 3 (15) | 4 (10) |
| Use of preoperative opioids — no. (%) | 9 (45) | 7 (35) | 16 (40) |
| Preoperative opioids — MME per day ¶ |  |  |  |
| Mean | 9.1±16.4 | 10.5±27.7 | 9.8±22.4 |
| Median | 0 | 0 | 0 |
| Use of intraoperative opioid infusion — no. (%) | 5 (25) | 7 (35) | 12 (30) |
| Length of surgery — min | 244±121 | 269±109 | 256±114 |

\* Plus-minus values represent means±SD. † American Society of Anesthesiologists (ASA) Physical Status
Classification ranges from I to VI, with higher values indicating greater number and severity of pre-anesthesia medical comorbidities. ‡ Total scores on the Charlson Comorbidity Index (CCI) range from 0 to 37, with higher scores indicating greater number and severity of pre-anesthesia medical comorbidities. § Inhaled nitrous oxide concentration of at least 50% for at least 1 continuous hour. ¶ Milligram morphine equivalents (MME) represent potencies of opioids relative to oral morphine, with higher values indicating higher opioid doses.

### OUTCOMES

For the primary outcome on post-infusion days 1, 2, and 3, the mixed-effects model (**Table 3**) showed no evidence of effect of group assignment on MADRS scores (95% CI −13.3 to 1.64, p=0.13, n=20 per arm). The MADRS rate of change also did not differ between groups (95% CI −1.54 to 4.93, p=0.30). An alternative model using change from pre-infusion baseline MADRS scores on the day of surgery (“day 0”) also showed no between-groups difference in change scores (95% CI −6.25 to 7.89, p=0.82). The rate of change for the MADRS change scores also did not differ between groups (95% CI −1.96 to 4.62, p=0.43). Missing MADRS scores among enrolled participants did not exceed 5% at any visit; therefore, missing data were not imputed for the primary outcome.

**Table 3.**
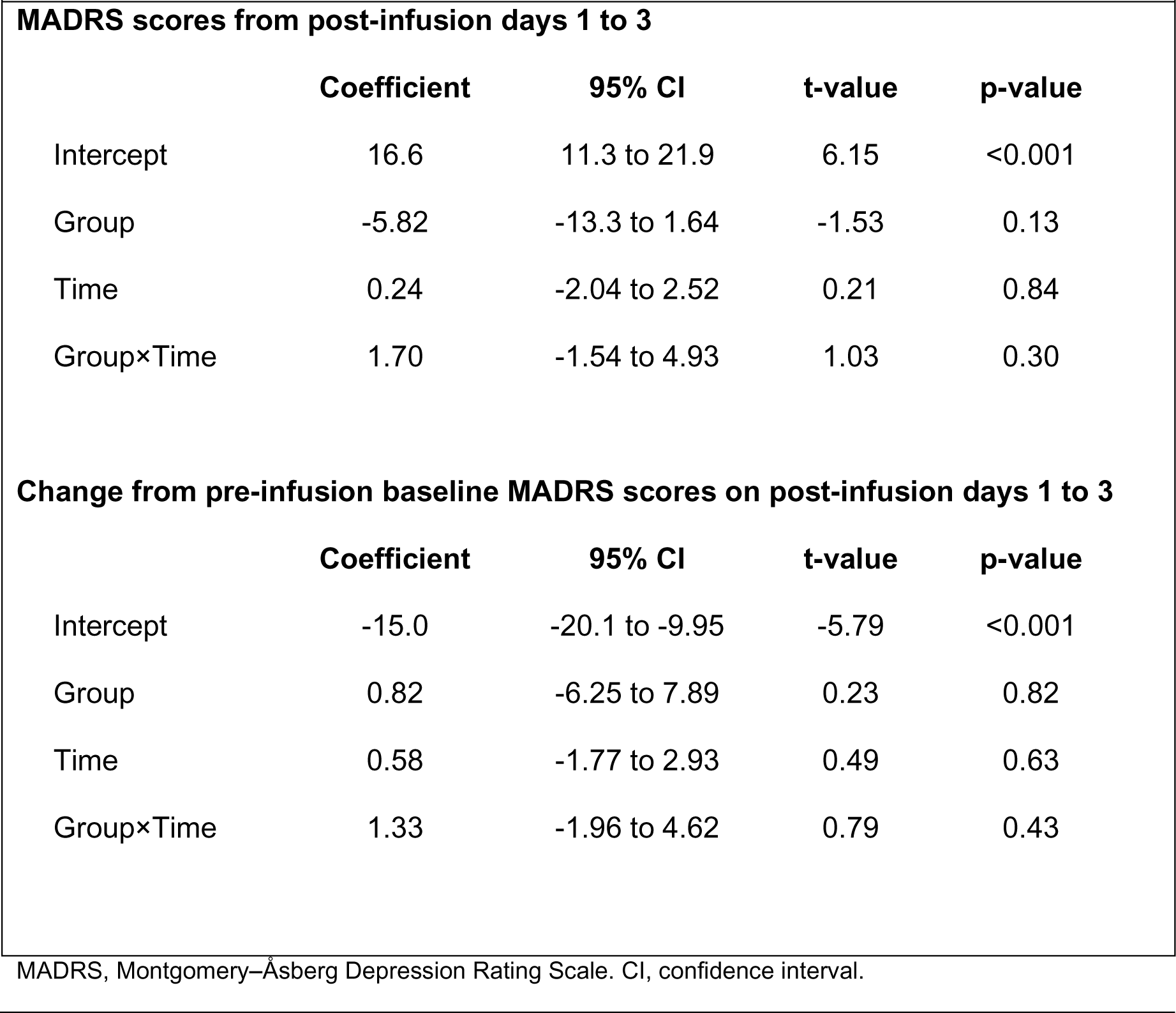
Linear Mixed Model Estimates for MADRS Scores.

Pre-infusion baseline MADRS scores on day 0 did not differ between trial groups (ketamine: 25.1 [SD 8.3]; placebo: 29.9 [SD 7.0]). From day 0 to day 1, the average change in MADRS scores was −12.4 points (SD 9.2) in the ketamine group and −14.7 points (SD 9.0) in the placebo group, corresponding to a mean decrease of 46% and 48% on the MADRS, respectively. In both groups, MADRS scores increased slightly on day 2 relative to the nadir at day 1, but the decrease from baseline persisted through all follow-up time points up to day 14 (**Figure 2A**). The HADS scores also followed a trajectory similar to the MADRS scores (**Extended Data Table 4**).

**Figure 2.**
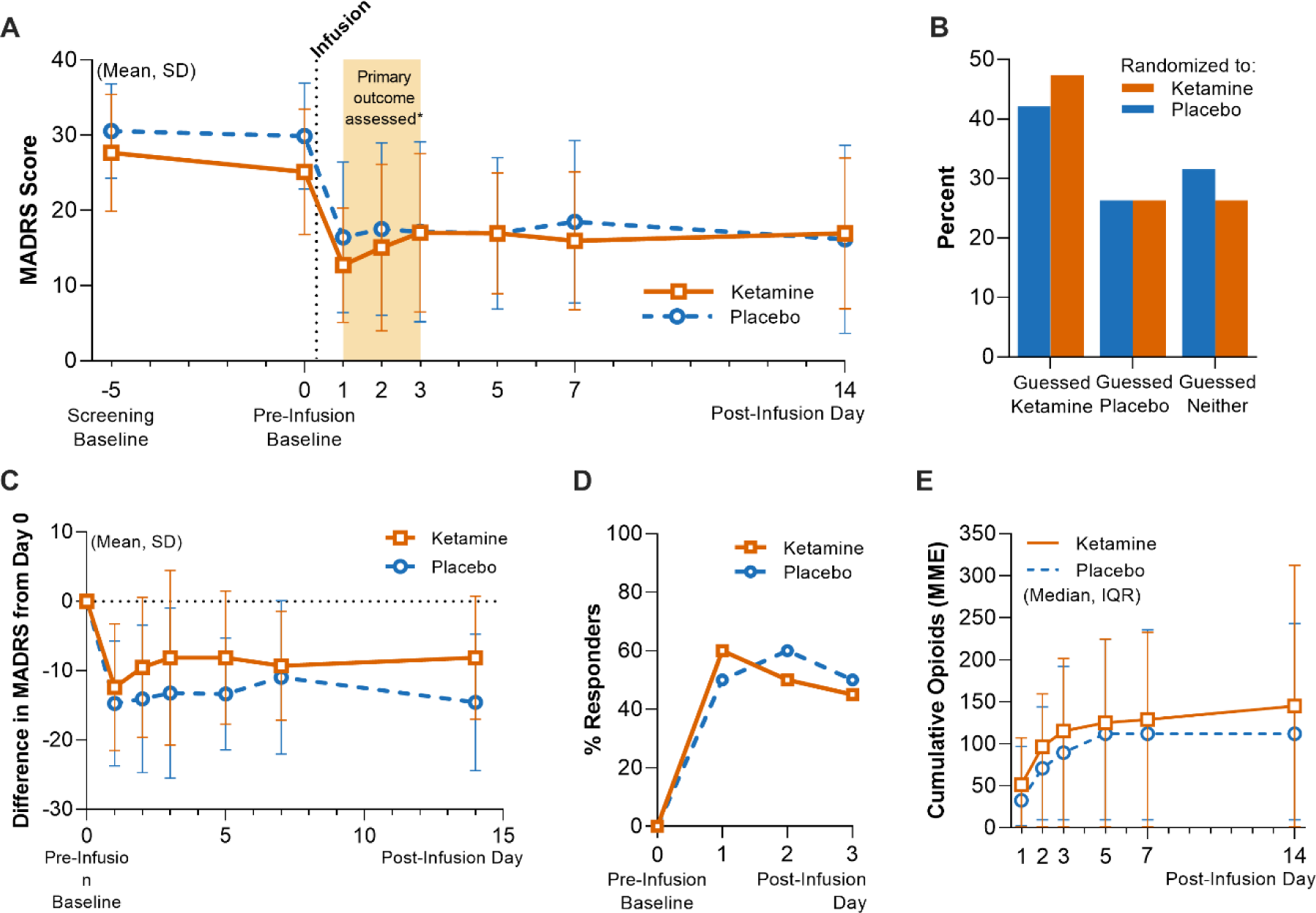
Depression Severity, Masking Assessment and Other Outcomes. Panel A shows the mean and standard deviation (SD) scores by group on the Montgomery-Åsberg Depression Rating Scale (MADRS; scores range from 0 to 60, with higher scores indicating greater depression); the screening baseline visit occurred on average 5 days before infusion on day 0. Panel B shows the distribution of guesses (as a percentage of each group; n=19 per group) made by participants when asked to guess which treatment they received after the last follow-up visit. Panel C shows the difference in MADRS scores relative to pre-infusion baseline scores obtained on day 0. Panels D shows the proportions of clinical response, respectively (as a percentage of each group; n=20 per group), within the first 3 days. Panel E shows the cumulative opioid consumption in MME by group, represented as median and interquartile range; both inpatient and outpatient opioids were included in the total.

At the end of the follow-up period (day 14), participants were asked to guess which intervention they had received; 36.8% of all participants guessed correctly, and the distribution of guesses between groups was comparable, with Cohen’s kappa = 0.33, indicating fair agreement between groups (**Figure 2B**). We performed an exploratory analysis to determine whether patients’ guesses regarding their treatment allocation were related to their final MADRS score (day 14). We regrouped MADRS data according to patient guess (“ketamine”, “placebo”, or “I don’t know”; **Extended Data Figure 1**). At day 14, aggregating across actual treatment allocations, mean [SD] MADRS scores were, for patients guessing “ketamine” (n=16), 10.1 [7.2]; guessing “placebo” (n=10), 19.2 [7.7]; replying “I don’t know” (n=12), 23.0 [13.7]. A simple logistic regression of day 14 MADRS score onto guessing either “ketamine” (coded ‘1’) or otherwise (coded ‘0’) suggested a significant inverse relationship between these two variables (odds ratio = 0.89 [95% CI 0.81 to 0.96]; p=0.001). MADRS change scores relative to day 0 are visualized in **Figure 2C**. Our secondary outcome was clinical response, defined as ≥50% reduction in MADRS scores from screening baseline. On post-infusion day 1, 60% and 50% of participants in the ketamine and placebo group, respectively, met criteria for clinical response. Rates of clinical response in both trial groups remained similar to each other on post-infusion days 2 and 3 (**Figure 2D**, **Table 4**). We also analyzed remission, which we defined in our study as MADRS score of ≤12. Remission occurred in 50% and 35% of participants in the ketamine and placebo group, respectively, on post-infusion day 1; this difference closed on post-infusion day 3, when 40% of both groups remained in remission (**Table 4**).

Cumulative opioid consumption did not differ between groups (**Figure 2E**). Of note, no participants received either preoperative or postoperative methadone or buprenorphine, nor were any maintained on opioid antagonist therapy. Average pain intensity on the BPI-SF at 14 days post-infusion was not different between groups (ketamine: 4.8 [SD 1.5]; placebo: 3.7 [SD 2.0]). Pain interference on the BPI-SF at day 14 post-infusion also did not differ between groups (ketamine: 6.2 [SD 2.2]; placebo: 5.7 [SD 3.5]). Hospital length of stay was longer in the placebo group (mean 1.9 [SD 1.7] days versus 4.0 [SD 3.3] days, p=0.02).

### PROTOCOL VIOLATIONS AND SAFETY

One participant in the ketamine group initially met inclusion criteria at screening but was retrospectively found not to have maintained her symptom severity on the morning of surgery (additive MADRS and HADS score of 30, below the minimum of 31). This participant was randomized and included in the ITT analysis. No protocol deviations related to the administration of the study drug occurred in this trial.

Adverse events were evaluated at every visit. Notably, one death in the ketamine group occurred 5 days post-infusion, which triggered the unmasking of treatment assignment. The patient had been discharged from the hospital on postoperative day 3, with no surgical or anesthetic complications documented prior to discharge. Subsequently, the patient experienced a witnessed cardiac arrest at home; advanced cardiac life support was initiated by paramedics and continued in the emergency room until the patient expired. One patient in the placebo group experienced a surgical complication requiring reoperation on postoperative day 3.

### SENSITIVITY ANALYSES

We tested whether our results were sensitive to: 1) a possible difference in pre-infusion baseline MADRS scores, and 2) the exclusion of participants who received nitrous oxide, an anesthetic which may also have antidepressant qualities^22^. We adjusted for a possible difference in pre-infusion baseline MADRS scores by including it as a fixed covariate and specifying random effects only for slopes in our mixed-effects model; this showed no between-group difference in MADRS scores (95% CI −8.41 to 4.49). When we applied our original mixed-effects model for the primary outcome after excluding 3 participants who received nitrous oxide at a concentration of 50% for at least 1 hour, there was also no between-group difference in MADRS scores (95% CI −13.3 to 2.74, p=0.13).

## Discussion

This randomized, triple-masked trial compared the short-term antidepressant efficacy of ketamine with placebo in adults with moderate-to-severe depression. There was no effect of treatment on our primary outcome, MADRS scores on days 1, 2, and 3 post-infusion. Baseline MADRS scores obtained at screening and on the day of surgery did not meaningfully differ between groups, supporting the effectiveness of randomization. In both trial groups, the observed decrease in MADRS score at day 1 was similar to, or exceeded, the decreases observed in previous ketamine trials in awake patients^29, 33–35^. The variance in MADRS change scores observed in our study is also comparable to previous studies in awake patients^19^, supporting our *a priori* power calculation to detect a between-group difference. The HADS, an alternative patient-rated depression scale, yielded a similar score trajectory as the MADRS and strengthens our conclusion that ketamine and placebo did not differentially impact mood in this trial.

Participant retention was excellent, with no loss to follow-up occurring within the primary outcome window. Notably, one participant death occurred in the ketamine group. However, this severe adverse event was attributed to underlying cardiovascular comorbidities rather than a direct result of trial procedures, consistent with previous analyses of cardiovascular safety outcomes after intravenous ketamine infusion^36^.

Both ketamine and placebo groups appeared to show a strong antidepressant response, though counter to our hypothesis, the magnitude of this response did not differ between groups. We review the available evidence for several potential interpretations of this result, while noting that the relatively small sample size, the unusual background of surgical anesthesia on which treatments were delivered, and our two-arm study design limit broadly generalizable conclusions about ketamine’s efficacy or mechanism. To the extent that the treatment effect in the ketamine group is similar to other studies, our data raise the possibility that antidepressant effects of ketamine may be achieved in the absence of a typical ketamine-induced conscious dissociative experience. However, a conscious dissociative experience may yield an even more robust response, significantly greater than that seen with placebo, in a situation where treatment-arm masking is maintained without the use of anesthesia. Furthermore, adjunctive psychotherapy may act syngergistically with the acute subjective effects of ketamine treatment (e.g. Ketamine-Assisted Psychotherapy), potentially outperforming ketamine-only treatment^37^, though controlled data on this form of ketamine therapy is still lacking^38^. Nonetheless, our successful masking of treatment allocation, and associated large apparent treatment effect across groups, may have implications for interpreting results from studies of acutely psychoactive treatments where trials are not designed for adequate treatment masking. In this study, patients’ guess regarding group allocation was strongly related with their MADRS score at the end of the study (14 days post infusion), potentially reflecting a prior belief about the efficacy of ketamine treatment for depression. Together, these data point to a major role for extra-pharmacological effects in the response to ketamine among depressed patients.

The surprisingly robust clinical response and remission rate observed in both arms of this trial raises the question of whether anesthetics besides ketamine may have antidepressant effects. N_2_O has been shown to improve depression symptoms in patients with TRD^22, 39^. However, only 3/40 participants in our study were exposed to N_2_O at a concentration of 50% for **≥** 1 hour, making it unlikely to impact depression scores at the group level, as confirmed in our sensitivity analyses. Propofol infusions and inhaled isoflurane have also shown antidepressant properties when given at doses that suppress EEG activity (“burst suppression”) for 15 minutes, over multiple administrations^40, 41^, though these findings are not consistent^42^ and differ substantially in depth and timing from standard surgical anesthesia used in our study (the recommended PSI range of 25-50 avoids burst suppression). We also considered the possibility that surgery and general anesthesia without ketamine has an antidepressant effect. However, our review of prior studies measuring symptoms of depression in the perioperative period strongly suggests otherwise. In the control arms (surgery and anesthesia alone) from studies in both depressed^43–47^ and non-depressed^48, 49^ patient populations, primary mood outcomes in the first days after surgery reflect either no significant change^43, 46, 48, 50, 51^ or possibly worsened symptoms^44, 49^. The anesthetic regimens used across these studies broadly resemble our own (most commonly propofol, sevoflurane and opioid-based), though the anesthetic depth was rarely noted. Taken together, these data suggest that non-pharmacological factors can strongly influence reported depression outcomes.

The lack of separation between placebo and ketamine groups in our trial may indicate that anesthetic agents blocked the antidepressant effects of ketamine. This possibility is somewhat difficult to reconcile with our observed effect size (comparing pre- to post-treatment), which is within the range of most previously observed ketamine effects sizes^6^. However, anesthetic agents may have interfered with neural mechanisms required for the antidepressant effect of ketamine, leaving in place, for example, large expectancy-driven antidepressant effects. Among several potential molecular and neural circuit-based mechanisms for the antidepressant effects of ketamine^52–54^, a popular model is that ketamine, via antagonism of cortical N-methyl-D-aspartate receptors (NMDARs), enhances cortical glutamate release and triggers neuroplastic changes through activity dependent release of neurotrophic factors. In this model, anesthetic agents could blunt ketamine effects by reducing excitability of cortical neurons. GABAergic anesthetic agents do indeed reduce cortical activity^55^, however emerging human data suggest that ketamine’s mechanisms may be substantially more complex than once thought. Based on a glutamate modulation theory of ketamine antidepressant action, multiple compounds have been tested as antidepressants in clinical trials and have not separated from placebo^56–61^, suggesting that other molecular targets may be involved. Further, interactions between ketamine and anesthetic agents like propofol may have both antagonistic and additive effects in cortical and subcortical networks^62, 63^, possibly linked to biologically and clinically significant actions of ketamine at, among other targets^1^, hyperpolarization-activated cyclic nucleotide-gated potassium channel 1 (HCN1)^62–66^, as well as opioid receptors^2, 67–70^. Opioids are also routinely used during surgical anesthesia, and recent evidence shows that blocking opioid receptors attenuates the antidepressant effect of ketamine^67, 71^. No patients were on opioid antagonist or partial agonist therapy, and the average daily MME use in both groups prior to surgery was relatively low. Our review of available data suggests that surgical anesthesia does not have substantial intrinsic antidepressant efficacy, and while we cannot exclude the possibility that anesthetic agents interfered with the antidepressant effect of ketamine, the antidepressant mechanism of ketamine in humans is not well enough characterized to make such a determination at this time. We hypothesize that a minimal dose of anesthetic agent may allow for adequate treatment masking and minimal interference with putative ketamine antidepressant mechanisms.

Baseline heterogeneity in psychiatric characteristics could potentially explain the smaller-than-anticipated difference in post-treatment depression scores. Although clinical and sociodemographic characteristics were largely similar between trial groups, there was a notable difference in current MDD episode length—with the ketamine group having a longer median episode duration (38 months) compared to the placebo group (17 months). A longer current MDD episode may predict more treatment resistance to traditional antidepressants^72^. However, studies comparing characteristics of responders and non-responders to ketamine therapy have been mixed, with some studies showing that current MDD episode length impacts treatment response^73^ while other studies do not^74, 75^. We also cannot rule out the effect of surgical heterogeneity between groups, which we did not control for in our recruitment design; however, between-group differences in case counts did not exceed 3 for any surgical department, and intraoperative factors, including length of surgery and types of anesthetic used, did not meaningfully differ between groups.

Other studies have also evaluated the effect of ketamine on mood ratings in surgical patients^47^; however, numerous methodological limitations prevent direct comparison to studies of intravenous ketamine for depression in the psychiatric literature. Frequently, these trials were not conducted in a population likely to meet criteria for moderate-to-severe MDD^48, 49, 76–78^. In the Prevention of Delirium and Complications Associated with Surgical Treatments (PODCAST) study—the largest study to date comparing ketamine with saline in surgical patients—depression scores were analyzed as a secondary outcome from a study designed to evaluate the efficacy of ketamine for postoperative delirium in patients >60 years old. Notably, only 9.6% of participants met the PHQ-8 cutoff for moderate depression preoperatively, and no diagnostic data or clinician-rated scales were reported^44^. Among perioperative studies that recruited patients with at least mild-moderate symptoms of depression^43, 44, 46, 50^, comparison with psychiatric trials in conscious patients is complicated by the use of non-standard ketamine doses and methods of administration^43, 44, 50^, reliance on patient-reported scales versus clinician-administered scales outcome measures^43, 44^, and treatment masking was either not described or not assessed. Kudoh *et al.* and Liu *et al.* enrolled surgical patients with mild to moderate depression severity. A 2021 RCT testing ketamine during surgical anesthesia required moderate-to-severe depression (MADRS ≥ 22) for eligibility; however, these participants underwent intracranial tumor resection^46^, a population we excluded due to the possibility of mood and personality changes associated with cortical lesions and resections of such lesions^79, 80^.

A key strength of our trial was the evaluation of participant masking. At their last follow-up visit, patients in both groups allocated their guesses in similar proportions and fewer than half guessed correctly. The intervention was effectively masked—an uncommon finding among antidepressant trials with ketamine. Assessment of masking is also rare among RCTs involving ketamine and electroconvulsive therapy (ECT), an important antidepressant treatment delivered to briefly anesthetized patients. Most RCTs evaluating the effect of adjunctive ketamine on ECT outcomes have found no benefit of ketamine given at doses of 0.5 mg/kg or greater^81^. Of the RCTs reviewed by McGirr *et al*., only one reported on masking effectiveness^82^.

Outcome expectancy related to the stated intent of the trial may drive apparent treatment effects. Previous studies of ketamine in surgical patients generally find that when patients are recruited to test ketamine’s antidepressant effect as a primary outcome, depression scores decrease postoperatively. Conversely, patients in the PODCAST study, who were recruited to participate in a trial focused on reducing postoperative delirium, depression symptoms (a secondary outcome) worsened slightly in the postoperative period.

One limitation of our study is that we did not assess the blind of the anesthesiologists who administered the study drug. While it is possible that close inspection of the intraoperative processed EEG could reveal changes consistent with a 0.5 mg/kg subanesthetic ketamine infusion^83^, we specifically instructed the anesthesiologists to avoid altering the patient’s anesthetic in response to the processed EEG during drug infusion (barring large excursions in PSI that correlate with patient awareness). Nonetheless, we cannot exclude the possibility that anesthesiologists who guessed the patient’s treatment allocation may have altered their anesthetic in a way that influenced postoperative mood.

Our results suggest that when differential subject-expectancy bias is minimized with successful masking, the treatment effect size of ketamine is reduced considerably. However, a major limitation of our study is that we did not measure treatment expectancies prior to randomization. Therefore, we cannot definitively conclude that subject-expectancy bias mediates the causal relationship between effective masking and smaller treatment effect sizes. Regardless of the intervention being tested, subject expectations of a positive outcome—also known as hope—may drive large decreases in depression symptoms seen in antidepressant trials^84^. Our trial design cannot distinguish between a null-effect of ketamine for depression and an occlusion of ketamine’s antidepressant effect through a placebo-like mechanism maintained in the absence of unmasking.

## CONCLUSION

This trial utilized surgical anesthesia to successfully mask the allocation of a single antidepressant dose of ketamine or placebo in a sample of depressed patients and found that depression scores at 1, 2, and 3 days post-infusion did not differ between trial groups. Both groups improved similarly. Secondary and exploratory outcomes also found no evidence of benefit for ketamine over placebo. These findings differ from those of prior antidepressant trials with ketamine conducted without adequate masking, which find robust effects of ketamine. Confounding surgical and anesthetic factors in our study prevent a determination of whether ketamine, on its own, is an effective short-term treatment of MDD. However, our robust, masked placebo response suggests that previously reported large effect sizes for ketamine may reflect a degree of expectancy bias. While it is impractical to use surgical anesthesia for most placebo-controlled trials, future studies of novel antidepressants with acute psychoactive effects should make stronger efforts to mask treatment assignment to minimize the effects of subject-expectancy bias.

## Supporting information

Suppl Protocol SAP Versions and Revision Summary

## Data Availability

De-identified participant data, data dictionaries, study protocol, and statistical analysis plan are available upon request after publication of the primary results manuscript in a peer-reviewed journal. To gain access to trial data for scientific research purposes, qualified researchers must submit a brief research proposal and statistical analysis plan for review and approval, and execute a data sharing agreement. Please direct all inquiries and requests to.

## DATA AVAILABILITY

De-identified participant data, data dictionaries, study protocol, and statistical analysis plan are available upon request after publication of this manuscript. To gain access to trial data for scientific research purposes, qualified researchers must submit a research proposal and statistical analysis plan for review and approval, and execute a data sharing agreement. Please direct all inquiries and requests to.

## CODE AVAILABILITY

R code used for data analysis is available upon request after publication of this manuscript. To gain access to the code for scientific research purposes, qualified researchers must submit a code use proposal for review and approval. Please direct all inquiries and requests to.

## ACKNOWLEDGEMENTS

BDH received support for this trial from the Society for Neuroscience in Anesthesia and Critical Care. We acknowledge Ms. Kayla Pfaff (Ohio University Heritage College of Osteopathic Medicine, Medical Student, Athens, Ohio, USA) and Dr. Rasmus Thordstein MD (Lund University, Lund, Sweden) for assistance with contacting patients, and Dr. Vishweshwara Ramachandran MBBS, MBA (Stanford University School of Medicine, Stanford, CA, USA) for implementing the PHQ-2 survey into the Anesthesia Preoperative Evaluation Clinic electronic workflow at Stanford.

Statistical support was provided by Data Studio (Department of Biomedical Data Science, Stanford University School of Medicine, Stanford, California, USA), which is supported by the National Center For Advancing Translational Sciences of the National Institutes of Health under Award Number UL1TR003142.

Screening and outcomes data were entered into Stanford REDCap (version 13.4.10), a secure online data capture platform (http://redcap.stanford.edu) developed and operated by Stanford Medicine Research IT team. The REDCap platform services at Stanford are subsidized by a) Stanford School of Medicine Research Office, and b) the National Center for Research Resources and the National Center for Advancing Translational Sciences, National Institutes of Health, through grant UL1 TR001085.

## Extended Data

**Figure 1.**
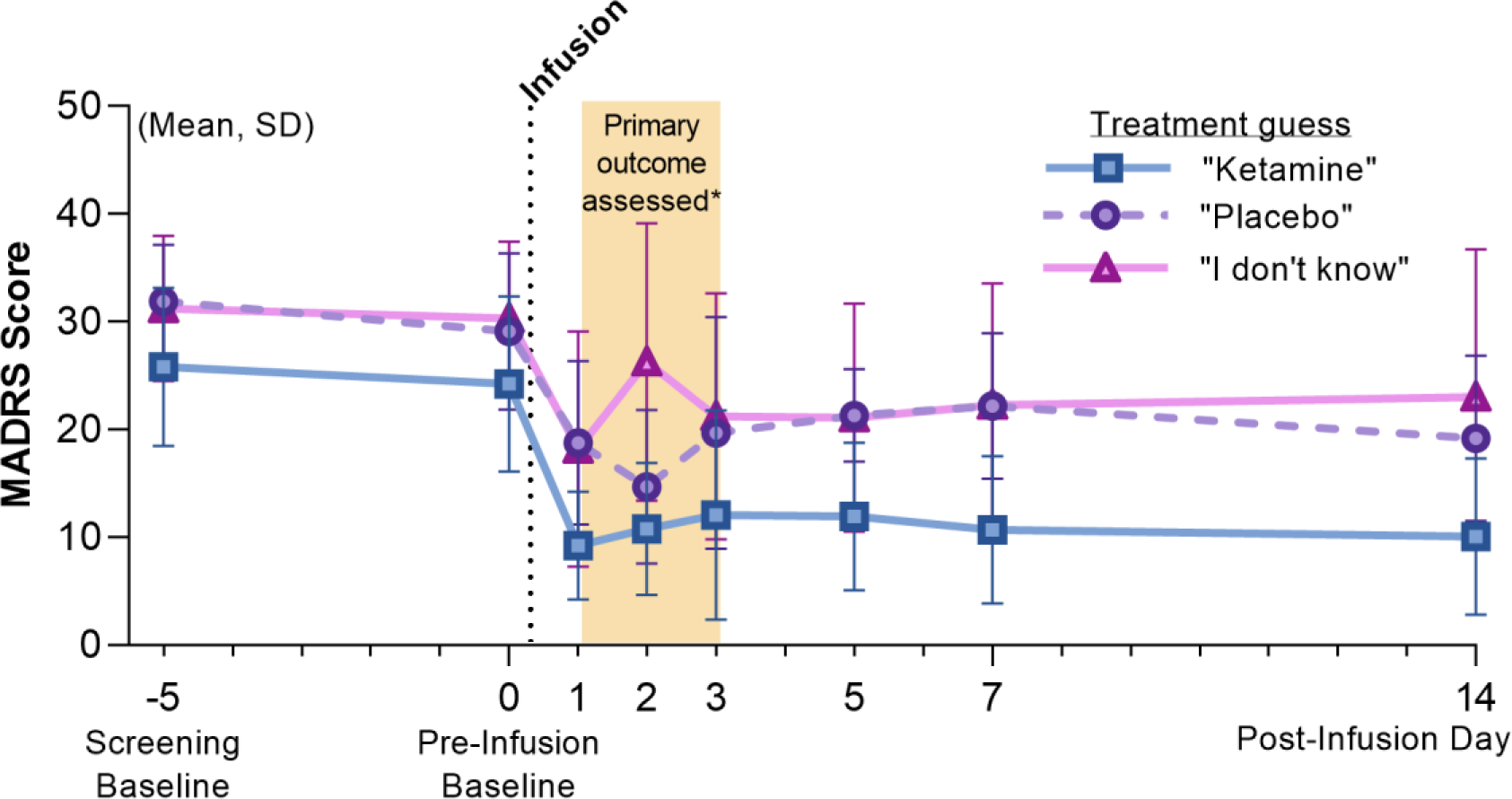
Depression ratings reanalyzed according to patient guess. On day 14, the final day of patient assessments, patients were asked the following questions: “What treatment do you think you received?” MADRS scores were reanalyzed according to their guess, irrespective of their true group allocation. Mean and standard deviation (SD) MADRS scores are shown using the alternate grouping: “Ketamine”, n=17; “Placebo”, n=10; “I don’t know”, n=11.

**Table 4.**
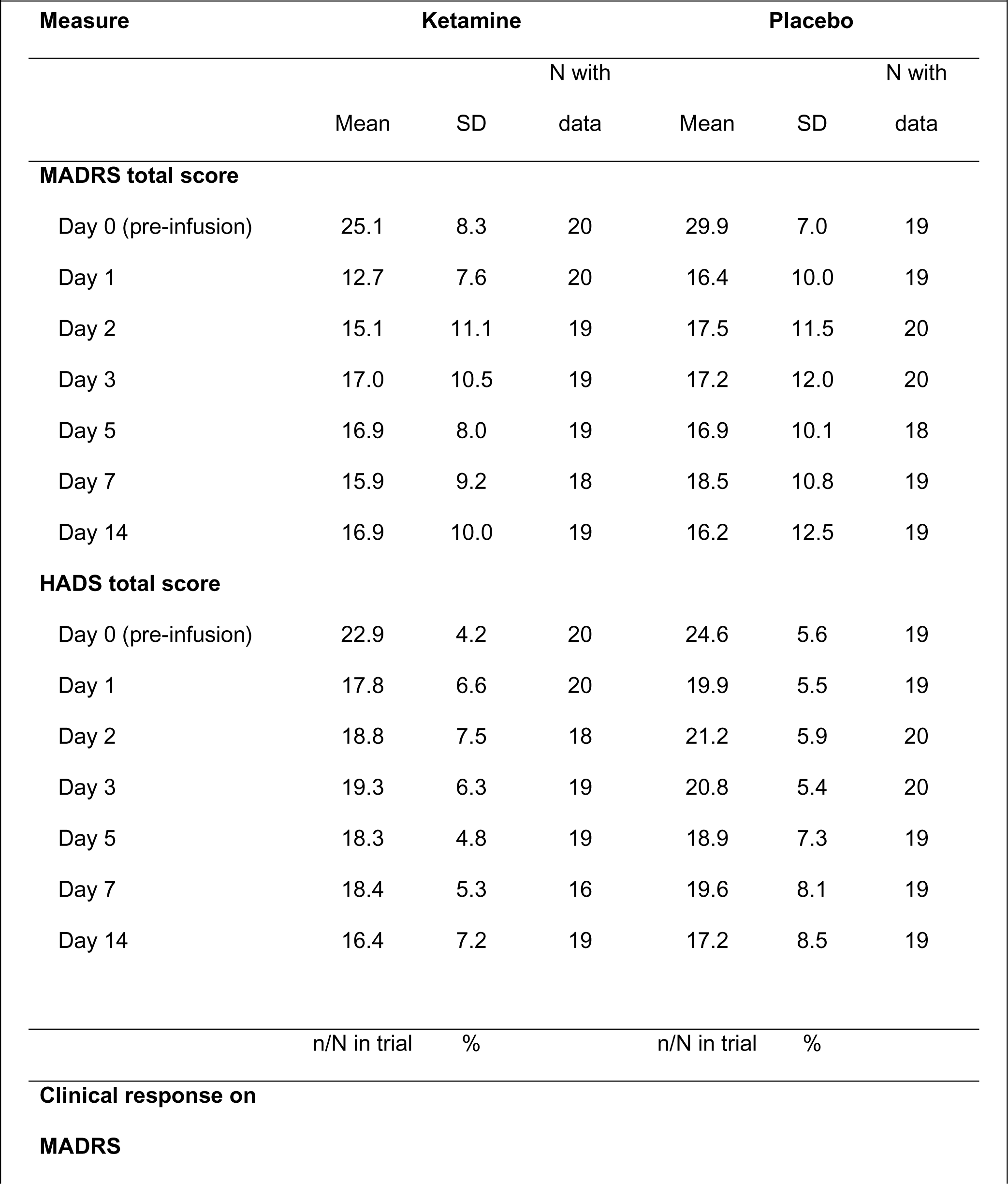
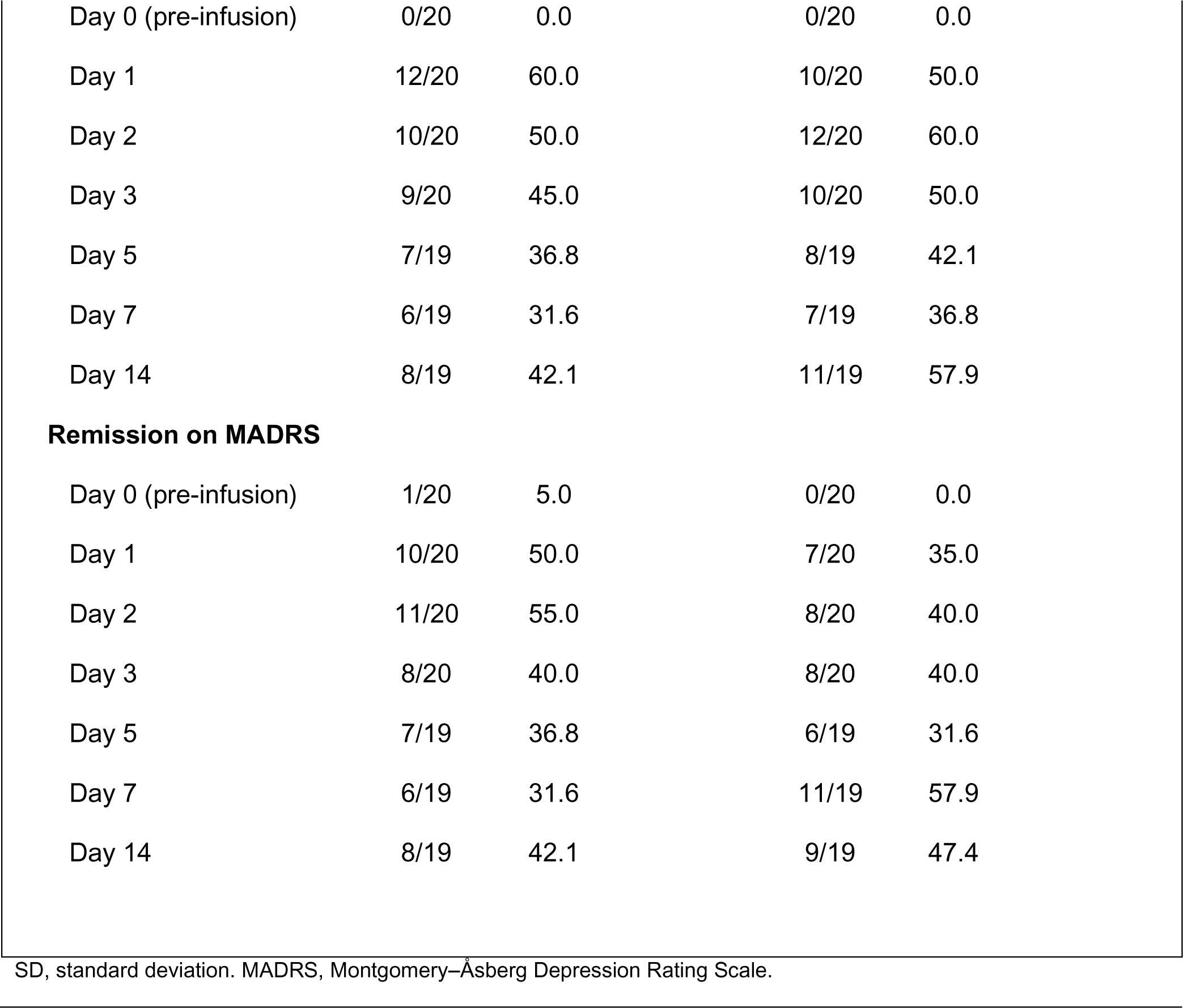
Depression Outcomes.

## Notes

### Competing Interest Statement

The authors have declared no competing interest.

### Clinical Trial

NCT03861988

### Funding Statement

This study received faculty start-up funding from the Department of Anesthesiology, Perioperative and Pain Medicine, Stanford University School of Medicine. BDH received support for this trial from the Society for Neuroscience in Anesthesia and Critical Care.

### Author Declarations

The Institutional Review Board of Stanford University gave ethical approval for this work

### Summary of Updates

Several modifications were made to this manuscript. Firstly, the word "Randomized" was added to the Title. The Abstract was revised to avoid implying a generalizable lack of ketamine efficacy based on our data. The Introduction was updated to include a secondary aim to examine whether a conscious dissociative reaction to ketamine is necessary for an antidepressant response. In the Trial Oversight section, the trial sponsors and the rationale for not employing a data safety monitoring board were provided. The Trial Design and Procedures section now includes a brief overview of the N=5 open-label pilot study and an explanation for the absence of anesthetic standardization in the main RCT. Additionally, logistic regression was incorporated into the Statistical Methods section as an exploratory analysis. The Participants section now includes enrollment dates for the first and last participants, as well as the last day of follow-up. The Outcomes section now includes the results of the exploratory logistic regression. In the Protocol Violations and Safety section, a description of a second severe adverse event unrelated to participation in the research study was added. In the Discussion, additional text was included to address whether surgery and anesthesia alone have an antidepressant effect. The section also summarizes prior research showing that anesthetics possess antidepressant properties at doses that suppress EEG activity and highlights the distinctions between the current study and prior perioperative studies that recruited patients with depression. The Conclusion was revised to emphasize limitations arising from confounding surgical and anesthetic factors. An acknowledgment of support for author BDH from the Society of Neuroscience in Anesthesia and Critical Care was added. A copy error in Table 2, which concerned the number and percentage of participants receiving isoflurane for surgical anesthesia, was corrected. An additional figure, Extended Data Figure 1, was included, illustrating a reanalysis of MADRS data based on participants' guesses at the end of the study. Lastly, additional minor events were added to the protocol revision summary table.

