## Supplementary material for "Randomized Trial of Ketamine Masked by Surgical Anesthesia in Depressed Patients": Suppl Protocol SAP Versions and Revision Summary

Stanford | School of Medicine
DEPARTMENT OF ANESTHESIOLOGY, PERIOPERATIVE AND PAIN MEDICINE

**Boris Dov Heifets, M.D., Ph.D.**
*Assistant Professor*
*Department of Anesthesiology, Perioperative and Pain Medicine*
*and by courtesy, Psychiatry and Behavioral Science*

Stanford University School of Medicine
1050 Arastradero Road, Building A, Rm. A151
Palo Alto, CA, 94304


June 9, 2023

Dear Reader,

This supplement contains the following items:

1. Original protocol (**p. 2-15**)
2. Final protocol (**p. 16-28**)
3. Summary of amendments to original protocol (**p. 29**)
4. Original statistical analysis plan (**p. 30-31**)
5. Final statistical analysis plan (**p. 32-44**)
6. Summary of amendments to the statistical analysis plan (**p. 45**)

Sincerely,

Boris Heifets M.D., Ph.D.

Original protocol (version 5, filed March 4, 2019)

### Stanford Placebo vs Intraoperative Ketamine Evaluation for Depression (SPIKED)

### STUDY PROTOCOL:
### Double-blind trial of intraoperative ketamine versus saline in patients with major depressive disorder undergoing anesthesia for total joint surgery

**1. NAME OF STUDY**

Double-blind trial of intraoperative ketamine versus saline in patients with major depressive disorder undergoing anesthesia for total joint surgery

**2. INVESTIGATORS:**

Principle Investigator: Boris Heifets, MD, PhD
Sub-Investigators: Alan Schatzberg, MD, Nolan Williams, MD, Theresa Lii, MD, Christopher Painter, MD, and William Maloney, MD

**3. SOURCE OF INSTITUTIONAL FUNDING:**

Departmental funds from the Stanford Department of Anesthesiology, Perioperative and Pain Medicine. This study is not industry sponsored. PTA # 1095509-2390-DAAAO

**4. SOURCES FROM WHOM WE ARE SEEKING FUNDS:**

None at the moment. Pending the results of this trial we will seek NIH or foundation grants.

**5. AIMS AND HYPOTHESIS**

**A. Aims:**

To determine whether intravenous ketamine, when used for its FDA-approved indication to supplement anesthesia, is associated with a measurable antidepressant effect in severely depressed patients undergoing total joint surgery when compared to placebo (normal saline infusion).

**B. Primary Outcome:**

Superiority of ketamine to saline infusion will be demonstrated by a statistically significant greater decrease in the composite score of the Hospital Anxiety and Depression Scale (HADS) and Montgomery and Asberg Depression Rating Scale (MADRS) when comparing baseline, pre-infusion scores to postoperative scores.

**C. Secondary Outcomes:**

- Pain at surgical site 0 - measured once between days 1 and 5 post procedure and again at 14 days post procedure
- Cumulative opioid consumption from postoperative day 0 up to 14
- Rates of intraoperative and postoperative complications/adverse events
- Hospital length of stay
- Metabolic profiles of patient breath samples

#### 6. GENERAL BACKGROUND

Major Depressive Disorder (MDD) is widely prevalent among patients preparing to have surgery, and it is a known risk factor for complications after surgery, including wound infection, myocardial infarction and opioid use disorder. Ketamine has emerged as an effective, rapid-acting antidepressant therapy for patients with MDD, and may be a useful tool to prevent MDD-related morbidity in the perioperative period. Ketamine has been well studied for MDD in outpatient clinics where it is given as an infusion (0.5 mg/kg over 40 minutes) in awake patients. Ketamine is often used as part of an anesthetic cocktail in sedated or anesthetized patients, but it is unknown whether ketamine has an antidepressant effect in this context.

We hope to discover whether ketamine has antidepressant efficacy in patients with severe MDD when given as an anesthetic adjunct. If ketamine is an effective antidepressant in this population under anesthesia, its use could be incorporated into a set of interventions to minimize the perioperative complications associated with MDD. While it is more efficient to deliver ketamine therapy intraoperatively, if this study finds that ketamine is ineffective in this setting, that result establishes a rationale to test treatment prior to the surgical encounter. We will also have gained important information on ketamine's antidepressant mechanism (e.g. it is blocked by other anesthetics, or it requires that the patient be conscious).

Only one published study addresses whether ketamine given with surgical anesthesia has any antidepressant effect in depressed patients (Kudoh et al., 2002). This study found a small antidepressant effect of questionable clinical significance, possibly due to studying patients with mild symptoms. This work, while promising, does not address whether ketamine has the profound antidepressant effect that has been repeatedly documented in nonsurgical psychiatric patients with severe MDD. Our proposed study specifically tests ketamine's antidepressant efficacy in this high risk group of patients with severe MDD that may particularly stand to benefit from remission of their depressive symptoms in the perioperative period. Other previous trials involving ketamine as an adjunct to anesthesia have uniformly been conducted in non-depressed patients and have limited relevance to our research question.

#### 7. PRELIMINARY UNPUBLISHED DATA

None at present. We have included an open-label, feasibility study as part of this protocol. However, there are already many published studies evaluating the antidepressant effect of ketamine at the dose used in our study, including one by the principal investigator and co-investigators (Williams *et al*., 2018).

#### 8. EXPERIMENTAL DESIGN, SCREENING, STUDY PROCEDURES AND DATA ANALYSIS

##### A. Design

We will first conduct an open-label, feasibility study involving 5 participants to identify which postoperative day to measure our primary endpoint in order to ensure that patients can cooperate with assessment interviews. **Once we have determined when to measure our primary endpoint, we will modify our registered trial (NCT03861988) on ClinicalTrials.gov**

**and submit to the IRB our updated study protocol for a placebo-controlled, double-blind, randomized controlled trial.**

After registering the trial and updating our protocol, we will conduct a single-center, double-blind, randomized, placebo-controlled trial involving a total of 40 participants (20 per group).

#### B. Study Population: Inclusion and Exclusion Criteria

Inclusion Criteria:
A subject will be eligible for inclusion when all of the following criteria are met:

1. Male or female, 18 to 80 years of age, inclusive, at screen.
2. Able to read, understand, and provide written, dated informed consent prior to screening. Participants will be deemed likely to comply with study protocol and communicate with study personnel about adverse events and other clinically important information.
3. Diagnosed with Major Depressive Disorder (MDD), single or recurrent, and currently experiencing a Major Depressive Episode (MDE) of at least eight weeks in duration, prior to screening, according to the criteria defined in the Diagnosis and Statistical Manual of Mental Disorders, Fifth Edition. The diagnosis of MDD will be made by a trained study staff member and supported by the Mini International Neuropsychiatric Interview - Module A.
4. Meet the threshold on the total combined HADS-MADRS score of >/=31 at both screening and day of surgery visits.
5. In sufficiently good health to proceed with planned orthopedic surgery, as ascertained by a standard preoperative clinic evaluation which includes medical history, physical examination (PE), clinical laboratory evaluations, any indicated cardiac testing, and final clearance by the attending anesthesiologists on the day of surgery.
6. Body mass index between 17-35kg/m2.
7. Concurrent psychotherapy will be allowed if the type (e.g., supportive, cognitive behavioral, insight-oriented, et al) and frequency (e.g., weekly or monthly) of the therapy has been stable for at least three months prior to screening and if the type and frequency of the therapy is expected to remain stable during the course of the subject's participation in the study.
8. Concurrent antidepressant therapy (e.g. SSRI or SNRI) and/or hypnotic therapy (e.g. zolpidem, zaleplon, melatonin, or trazodone) will be allowed if the therapy has been stable for at least 4 weeks prior to screening and if it is expected to remain stable during the course of the subject's participation in the study.

Exclusion Criteria:
A potential participant will NOT be eligible for participation in this study if any of the following criteria are met:

1. Female that is pregnant or breastfeeding. These women would not be candidates for elective total joint replacement under any circumstances, and therefore would be screened out from our study population at their routine preoperative evaluation. Women of childbearing potential are routinely screened for pregnancy at their preoperative visit by urine hCG testing if clinically indicated.

2. Total HADS-MADRS score of <31 at either the screening or day of surgery visits.
3. Current diagnosis of a Substance Use Disorder (Abuse or Dependence, as defined by DSM-V), with the exception of nicotine dependence, at screening or within six months prior to screening. The diagnosis of Substance Abuse Disorder will be made by a trained study staff member and supported by the Mini International Neuropsychiatric Interview – Module J.
4. History of schizophrenia or schizoaffective disorders, or any history of psychotic symptoms in the current or previous depressive episodes.
5. In the judgment of the investigator, the subject is at significant risk for suicidal behavior during the course of his/her participation in the study.
6. Has dementia, delirium, amnestic, or any other cognitive disorder.
7. Has a clinically significant abnormality on the screening physical examination that would otherwise preclude the patient from having surgery.
8. Participation in any clinical trial with an investigational drug or device within the past month or concurrent to study participation.
9. Lifetime history of surgical procedures involving the brain or meninges, encephalitis, meningitis, degenerative central nervous system disorder (e.g., Alzheimer's or Parkinson's Disease), epilepsy, mental retardation, or any other disease/procedure/accident/intervention associated with significant injury to or malfunction of the central nervous system (CNS), or a history of significant head trauma within the past two years.
10. Presents with any of the following lab abnormalities w/in the past 6 months:

a. Thyroid stimulating hormone (TSH) outside of the normal limits and clinically significant as determined by the investigator. Free thyroxine (T4) levels may be measured if TSH level is high. Subject will be excluded if T4 level is clinically significant.
b. Any other clinically significant abnormal laboratory result at the time of the screening exam.

11. History of hypothyroidism and has been on a stable dosage of thyroid replacement medication for less than six months prior to screening.(Subjects on a stable dosage of thyroid replacement medication for at least six months or more prior to screening are eligible for enrollment.)
12. History of hyperthyroidism which was treated (medically or surgically) less than six months prior to screening.
13. Any current or past history of any physical condition which in the investigator's opinion might put the subject at risk or interfere with study results interpretation.
14. Patients currently maintained on high dose opioids (>90 morphine equivalents per day) prior to surgery.

#### C. Statisical Analysis

##### Sample Size

Based on published data from other investigators (Zarate *et al*., 2006) and our own published work (Williams *et al*., 2018), we estimate a sample size of 15 patients per group, alpha=0.05, power = 80% to detect a 30% change in depression rating from baseline. We will include an additional 5 patients per group to account for participant dropout, for a total of 20 patients per group.

##### Analysis Plan

1. Statistical analyses will be performed by the investigators using GraphPad Prism 8.

2. Baseline characteristics and demographics between groups will be compared using t-tests for continuous and ordinal variables and chi-square tests for categorical variables.
3. Primary outcome: superiority of ketamine to saline infusion will be demonstrated by a statistically significant greater decrease in the composite score of the Hospital Anxiety and Depression Scale (HADS) and Montgomery and Asberg Depression Rating Scale (MADRS) when comparing baseline, pre-infusion scores to postoperative scores, using a t-test ($p<0.05$, 2-sided, adjusting for multiple comparisons).
4. Secondary outcomes between groups will be compared using t-tests for continuous and ordinal variables and chi-squared tests for categorical variables.

#### D. Study Procedures from Screening to Closeout

Screening

Potential participants will be identified at the time of their initial orthopedic clinic visit for total joint arthroplasty.

Eligible patients will be identified by clinical coordinator or study physician based on a short questionnaire response included with their orthopedic surgery paperwork. Eligibility questionnaires and related consent procedures are obtained through a collaboration with an ongoing IRB-approved study (PI Dr. Jennifer Hah, IRB #43163), and not through this protocol.

Eligibility will be determined by the Patient Health Questionnaire 8-item (PHQ-8, which is the same as PHQ-9 but omitting a question about suicide) included with orthopedic clinic paperwork. Patients are considered eligible for the study if their PHQ-8 score is 12 or higher (indicating moderate to severe depression). Suicidal ideation will be assessed and managed as described in Safety Plan for Patients Who Endorse Active Suicidality. A study nurse/coordinator or study physician (“study staff member”) will perform a chart review to verify eligibility and contact the patient in person to inform them about the study. A study staff member will subsequently obtain consent in-person prior to any additional screening, and they will ensure the patient has capacity to consent. Consent will be obtained at either their preoperative orthopedic visit, at a separate visit to Stanford Hospital, or on the morning of surgery prior to administration of any medication. **Informed consent will be obtained prior to full evaluation of inclusion and exclusion criteria.**

After providing informed consent, the participant will be offered the option to complete screening in the following ways:

1. During a preoperative visit to Stanford Medicine Outpatient Center.
2. By phone or video interview (using Zoom, which is PHI compliant) prior to surgery.
3. In person visit to Stanford Medical Center.
4. On the day of surgery, prior to surgery at Stanford Hospital; patients will be asked to check in 30-45 minutes early to allow time for screening.

Participants will have the standard preoperative workup and management for total joint arthroplasty, including diagnostic labs/studies and perioperative medication adjustment as needed.

During screening, investigators must document a patient history of Major Depressive Disorder (MDD), single or recurrent. The patient must be currently experiencing a Major Depressive Episode (MDE) of at least eight weeks in duration prior to screening, according to the criteria defined in the Diagnosis and Statistical Manual of Mental Disorders, Fifth Edition. The diagnosis of MDD will be made by a trained study staff member and supported by the Mini International Neuropsychiatric Interview (MINI) - Module A. Patients with a current diagnosis of Alcohol or Substance Use Disorder (Abuse or Dependence, as defined by DSM-V), with the exception of nicotine dependence, at screening or within six months prior to screening will be excluded. The diagnosis of Alcohol or Substance Abuse Disorder will be made by a trained study staff member and supported by the Mini International Neuropsychiatric Interview – Modules I and J.

Concurrent psychotherapy will be allowed if the type (e.g., supportive, cognitive behavioral, insight-oriented, etc.) and frequency (e.g., weekly or monthly) of the therapy has been stable for at least 3 months prior to screening and if the type and frequency of the therapy is expected to remain stable during the course of the subject's participation in the study.

Concurrent antidepressant therapy (e.g. SSRI or SNRI) and/or hypnotic therapy (e.g. zolpidem, zaleplon, melatonin, or trazodone) will be allowed if the therapy has been stable for at least 4 weeks prior to screening and if it is expected to remain stable during the course of the subject's participation in the study.

Once patients agree to participate in the study and have signed the informed consent document, the following screening procedures will be performed:

By Physician or Study Staff:
- Concomitant Medication Review
- Medical and Psychiatric History
- Review of preoperative evaluation data including physical exam, vital signs, any lab work (including a pregnancy test, if applicable) or cardiac testing performed in the course of standard preoperative medical clearance
- Urine toxicology screen test for drugs of abuse
- Mini International Neuropsychiatric Interview (MINI) - Modules A, I & J
- Columbia Suicidality Severity Rating Scale - short screen
- Montgomery-Asberg Depression Rating Scale (MADRS)
- Maudsley Staging Method (MSM)

By the Participant, reviewed by Investigators:
- Hospital Anxiety and Depression Scale (HADS)
- Brief Pain Inventory (BPI)

Participant Compensation:
- $50 after completion of Screening and $50 upon completion of study at day 14. If patients come to Stanford Hospital for the purpose of screening, they will be reimbursed an additional $50 for their time and transportation costs.

Safety Plan for Patients Who Endorse Active Suicidality

A patient may be identified to be at risk for suicidal behavior after consent and during our extended screening process. The Columbia Suicide Severity Rating Scale (CSSRS) – Short Screen is administered face to face, by phone, or by video call (Stanford Zoom) by a study staff member. A patient may be identified as “low”, “moderate” or “high” risk. For any patient scoring “moderate” or “high” risk, the study staff will immediately contact Dr. Boris Heifets or Ms. Robin Okada (RN and study coordinator).

If Dr. Heifets or Ms. Okada are notified of a patient at risk for suicidal behavior, Dr. Heifets will contact that patient, verify their response and decide if a wellness check from police is needed or other interventions. That patient will be offered immediate assistance, including calling an ambulance for transport to the Emergency Department, and their primary physician will be informed.

Randomization

Only Investigational Drug Services (IDS) Pharmacy will be aware of drug allocation prior to completion of data collection for the randomized, placebo-controlled study. A computer-generated randomization scheme will be used to randomize participants to receive either ketamine or saline (allocation ratio 1:1, blocks of 4). The study participants, intraoperative anesthesiologists, and study staff conducting postoperative assessments will be blinded to drug allocation until data collection is completed.

Infusions: The study intervention will occur during surgery (Study Day 0). Participants will be randomized to one of two groups: Group A (n=20) will receive a ketamine infusion of 0.5 mg/kg over 40 min during surgery, beginning after anesthetic induction with propofol. Group B (n=20) will receive a saline infusion over 40 min during surgery, beginning after anesthetic induction with propofol. Infusion (instead of bolus) dosing of ketamine was chosen since infusions have demonstrated antidepressant efficacy in outpatient psychiatric populations, as well as theoretically lower risk of adverse events due to lower peak serum concentrations compared to bolus dosing. A patient who is ***not*** enrolled in the study has a reasonable chance of receiving the same treatments (ketamine or saline) at comparable doses as those tested in the study.

Monitoring: Routine vital signs will be collected before and after surgery by the participant’s nurse and reviewed by investigators. Each participant will have continuous physiological monitoring during their anesthetic. A SedLine® Brain Function Monitor will be used to evaluate depth of anesthesia/sedation. If the participant experiences an adverse event temporally related to the infusion where corrective action by the intraoperative anesthesiologist requires unblinding, then the study infusion drug will be revealed to the anesthesiologist.

Breath Collection: Three breath samples will be collected on day of surgery (Study Day 0). In the preoperative holding area, the patient will be asked to exhale 1 breath into a Tedlar® breath bag for breath sample #1. During surgery, after completion of the study infusion, breath sample #2 will be collected by study staff or the intraoperative anesthesiologist. This sample will be obtained by connecting a syringe to the already accessible port of the ventilation circuit which is used to sample and measure end-tidal CO2 during surgery. The contents of the syringe will be then placed into a Tedlar® breath bag. Once the surgery is finished and the patient has recovered from anesthesia, the patient will be asked to exhale 1 breath into a Tedlar® breath bag for breath sample #3. All samples collected will be labeled with a unique identifier number. The patient's hospital course will be followed by their medical record. Breath samples will be analyzed by a gas mass spectrometer in a building adjacent to and on the same floor as the operating rooms in Grant S290. The analysis will occur on the same day of surgery.

Breath Collection Data Safety Monitoring Plan: The breath collection protocol is a low risk protocol which will not require a data safety monitoring plan. No therapeutic agent is being used and the sample collection is low risk similar to collecting a urine, blood or stool sample.

Data Collection Procedures

The first 5 patients that meet inclusion/exclusion criteria will be entered into the open-label feasibility study; all will receive a ketamine infusion. All other patients enrolled thereafter will be entered into the double-blind, randomized, placebo-controlled study. The day of surgery will be considered Day 0 of the study. Baseline assessments must be performed *before* the patient receives any psychoactive premedication (e.g. midazolam, fentanyl, gabapentin, oxycodone). Assessments will be performed either in the hospital or, if the patient has been discharged (typically on postoperative Day 2), by phone.

**Day 0 (Baseline Assessments):**
By Physician or Study Staff:
- Vital Signs
- Concomitant Medication Review
- Adverse Events Review
- Montgomery-Asberg Depression Rating Scale (MADRS)
- Collect breath sample #1

By the Participant, reviewed by Investigators:
- Hospital Anxiety and Depression Scale (HADS)
- Brief Pain Inventory (BPI)

**Day 0 (During Surgery):**
By the Intraoperative Anesthesiologist:
- Ketamine infusion or placebo (saline) infusion
- Continuous physiological monitoring
- SedLine® Brain Function Monitor to evaluate depth of anesthesia/sedation
- Collect breath sample #2, after infusion is complete

**Day 0 (Postoperative, Post-PACU):**
By Physician or Study Staff:
- Vital Signs
- Concomitant Medication Review
- Adverse Events Review
- Montgomery-Asberg Depression Rating Scale (MADRS)
- Collect breath sample #3

By the Participant, reviewed by Investigators:
- Hospital Anxiety and Depression Scale (HADS)
- Brief Pain Inventory (BPI)

**Day 1:**
By Physician or Study Staff:
- Vital Signs
- Concomitant Medication Review
- Adverse Events Review
- Montgomery-Asberg Depression Rating Scale (MADRS)

By the Participant, reviewed by Investigators:
- Hospital Anxiety and Depression Scale (HADS)
- Brief Pain Inventory (BPI)

**Day 2:**
By Physician or Study Staff:
- Vital Signs (inpatient only)
- Concomitant Medication Review
- Adverse Events Review
- Montgomery-Asberg Depression Rating Scale (MADRS)

By the Participant, reviewed by Investigators:
- Hospital Anxiety and Depression Scale (HADS)
- Brief Pain Inventory (BPI)

**Day 3 (inpatient or via phone):**
By Physician or Study Staff:
- Vital Signs (inpatient only)
- Concomitant Medication Review
- Adverse Events Review
- Montgomery-Asberg Depression Rating Scale (MADRS)

By the Participant, reviewed by Investigators:
- Hospital Anxiety and Depression Scale (HADS)
- Brief Pain Inventory (BPI)

**Day 5 (inpatient or via phone):**
By Physician or Study Staff:

- Vital Signs (inpatient only)
- Concomitant Medication Review
- Adverse Events Review
- Montgomery-Asberg Depression Rating Scale (MADRS)

By the Participant, reviewed by Investigators:
- Hospital Anxiety and Depression Scale (HADS)
- Brief Pain Inventory (BPI)

**Day 7 (inpatient or via phone):**
By Physician or Study Staff:
- Vital Signs (inpatient only)
- Concomitant Medication Review
- Adverse Events Review
- Montgomery-Asberg Depression Rating Scale (MADRS)

By the Participant, reviewed by Investigators:
- Hospital Anxiety and Depression Scale (HADS)
- Brief Pain Inventory (BPI)

**Day 14 (inpatient or via phone):**
By Physician or Study Staff:
- Ask participants what treatment they believe they received
- Reveal treatment to participant
- Vital Signs (inpatient only)
- Concomitant Medication Review
- Adverse Events Review
- Montgomery-Asberg Depression Rating Scale (MADRS)

By the Participant, reviewed by Investigators:
- Hospital Anxiety and Depression Scale (HADS)
- Brief Pain Inventory (BPI)

Participant Compensation:
- $50 after completion of all study assessments

Study Procedures Matrix

| | Screen-ing | Study Phase | | | | | | | | |
|---|---|---|---|---|---|---|---|---|---|---|
| | | Day 0 (Baseline) | Day 0 (Intraop) | Day 0 (Postop) | Day 1 | Day 2 | Day 3 | Day 5 | Day 7 | Day 14 |

| | | | | | | | | | | |
|---|---|---|---|---|---|---|---|---|---|---|
| PHQ-8 | X | | | | | | | | | |
| Inclusion/Exclusion Criteria | X | | | | | | | | | |
| Demographic data | X | | | | | | | | | |
| Consent | X | | | | | | | | | |
| Concomitant Medication Review | X | X | | X | X | X | X | X | X | X |
| Medical and Psychiatric History | X | | | | | | | | | |
| Physical Exam | X | | | | | | | | | |
| Vital Signs | X | X | | X | X | X | O | O | O | O |
| Continuous physiological monitoring | | | X | | | | | | | |
| Urine toxicology screen test | X | | | | | | | | | |
| Labs and studies as warranted by standard preoperative medical clearance (includes pregnancy test) | X | | | | | | | | | |
| Mini International Neuropsychiatric Interview (MINI) - A , I & J | X | | | | | | | | | |
| Hospital Anxiety and Depression Scale (HADS) | X | X | | X | X | X | X | X | X | X |
| Montgomery Asberg Depression Rating Scale (MADRS) | X | X | | X | X | X | X | X | X | X |
| Columbia Suicide Severity Rating Scale (CSSRS) – Short Screen | X | | | | | | | | | |

| | | | | | | | | | | |
|---|---|---|---|---|---|---|---|---|---|---|
| Maudsley Staging Method (MSM) | X | | | | | | | | | |
| Brief Pain Inventory | X | X | | X | X | X | X | X | X | X |
| Randomization | X | | | | | | | | | |
| Ketamine infusion or Placebo (saline) infusion | | | X | | | | | | | |
| SedLine® Brain Function Monitor | | | X | | | | | | | |
| Collect patient breath sample | | X | X | X | | | | | | |
| Ask participants what treatment they believe they received | | | | | | | | | | X |
| Treatment revealed to participant | | | | | | | | | | X |
| Participant compensation | X | | | | | | | | | X |

X – required
O - inpatient only

#### 9. SIGNIFICANCE

Major Depressive Disorder (MDD) is widely prevalent among patients preparing to have surgery, and is a known risk factor for complications after surgery, including wound infection, myocardial infarction and opioid use disorder. Ketamine has emerged as an effective, rapid-acting antidepressant therapy for patients with MDD, and may be a useful tool to prevent MDD-related morbidity in the perioperative period. Ketamine has been well studied for MDD in outpatient clinics where it is given as an infusion (0.5 mg/kg over 40 minutes) in awake patients. Ketamine is often used as part of an anesthetic cocktail in sedated or anesthetized patients, but it is unknown whether ketamine has an antidepressant effect in this context. We will determine whether a ketamine infusion, compared to placebo (normal saline infusion), has an antidepressant effect when given during surgical anesthesia. If ketamine is an effective antidepressant in this population under anesthesia, its use could be incorporated into a set of interventions to minimize the perioperative complications associated with MDD.

2. Final protocol (version 7, filed October 22, 2021)

### Stanford Placebo vs Intraoperative Ketamine Evaluation for Depression (SPIKED)

### STUDY PROTOCOL:
### Double-blind trial of intraoperative ketamine versus saline in patients with major depressive disorder undergoing non-cardiac surgery
### NCT 03861988

**1. NAME OF STUDY**
Double-blind trial of intraoperative ketamine versus saline in patients with major depressive disorder undergoing anesthesia for non-cardiac surgery

**2. INVESTIGATORS:**
Principle Investigator: Boris Heifets, MD, PhD
Sub-Investigators: Theresa Lii, MD, Alan Schatzberg, MD,

**3. SOURCE OF INSTITUTIONAL FUNDING:**
Departmental funds from the Stanford Department of Anesthesiology, Perioperative and Pain Medicine. This study is not industry sponsored.

**4. SOURCES FROM WHOM WE ARE SEEKING FUNDS:**
Pending the results of this trial we will seek NIH or foundation grants.

**5. AIMS AND HYPOTHESIS**

**A. Aims:**
To determine whether intravenous ketamine, when used for its FDA-approved indication to supplement anesthesia, is associated with a measurable antidepressant effect in severely depressed patients undergoing non-cardiac surgeries when compared to placebo (normal saline infusion).

**B. Primary Outcome:**
We plan to evaluate the antidepressant superiority of ketamine to placebo by assessing postoperative MADRS scores at multiple timepoints. MADRS (Montgomery-Asberg Depression Rating Scale) is a validated measure of depression severity used routinely in antidepressant trials.

**C. Secondary Outcomes:**
- Proportion of participants with clinical response (defined as a ≥50% reduction in MADRS score from baseline)
- Proportion of participants with remission (defined as a MADRS score of ≤12 on day 14)
- Hospital Anxiety and Depression Scale
- Cumulative inpatient opioid use
- Hospital length of stay
- Average inpatient opioid use per day
- Opioid use at postop day 7
- Opioid use at postop day 14
- Postoperative numeric pain scores
- Postoperative pain interference scores
- Immunological blood markers

**6. GENERAL BACKGROUND**

Major Depressive Disorder (MDD) is widely prevalent among patients preparing to have surgery, and it is a known risk factor for complications after surgery, including wound infection, myocardial infarction and opioid use disorder. Ketamine has emerged as an effective, rapid-acting antidepressant therapy for patients with MDD, and may be a useful tool to prevent MDD-related morbidity in the perioperative period. Ketamine has been well studied for MDD in outpatient clinics where it is given as an infusion (0.5 mg/kg over 40 minutes) in awake patients. Ketamine is often used as part of an anesthetic cocktail in sedated or anesthetized patients, but it is unknown whether ketamine has an antidepressant effect in this context.

We hope to discover whether ketamine has antidepressant efficacy in patients with severe MDD when given as an anesthetic adjunct. If ketamine is an effective antidepressant in this population under anesthesia, its use could be incorporated into a set of interventions to minimize the perioperative complications associated with MDD. While it is more efficient to deliver ketamine therapy intraoperatively, if this study finds that ketamine is ineffective in this setting, that result establishes a rationale to test treatment prior to the surgical encounter. We will also have gained important information on ketamine's antidepressant mechanism (e.g. it is blocked by other anesthetics, or it requires that the patient be conscious).

Only one published study addresses whether ketamine given with surgical anesthesia has any antidepressant effect in depressed patients (Kudoh et al., 2002). This study found a small antidepressant effect of questionable clinical significance, possibly due to studying patients with mild symptoms. This work, while promising, does not address whether ketamine has the profound antidepressant effect that has been repeatedly documented in nonsurgical psychiatric patients with severe MDD. Our proposed study specifically tests ketamine's antidepressant efficacy in this high risk group of patients with severe MDD that may particularly stand to benefit from remission of their depressive symptoms in the perioperative period. Other previous trials involving ketamine as an adjunct to anesthesia have uniformly been conducted in non-depressed patients and have limited relevance to our research question.

#### 7. PRELIMINARY UNPUBLISHED DATA

None at present. We have included an open-label, feasibility study as part of this protocol. However, there are already many published studies evaluating the antidepressant effect of ketamine at the dose used in our study, including one by the principal investigator and co-investigators (Williams et al., 2018).

#### 8. EXPERIMENTAL DESIGN, SCREENING, STUDY PROCEDURES AND DATA ANALYSIS

##### A. Design

We will first conduct an open-label, feasibility study involving 5 participants to identify which postoperative day to measure our primary endpoint in order to ensure that patients can cooperate with assessment interviews. **Once we have determined when to measure our primary endpoint, we will modify our registered trial (NCT03861988) on ClinicalTrials.gov and submit to the IRB our updated study protocol for a placebo-controlled, double-blind, randomized controlled trial.**

After registering the trial and updating our protocol, we will conduct a single-center, double-blind, randomized, placebo-controlled trial involving a total of 40 participants (20 per group).

##### B. Study Population: Inclusion and Exclusion Criteria

Inclusion Criteria:

A subject will be eligible for inclusion when all of the following criteria are met:

1. Male or female, 18 to 80 years of age, inclusive, at screen.
2. Able to read, understand, and provide written, dated informed consent prior to screening. Participants will be deemed likely to comply with study protocol and communicate with study personnel about adverse events and other clinically important information.
3. Diagnosed with Major Depressive Disorder (MDD), single or recurrent, and currently experiencing a Major Depressive Episode (MDE) of at least eight weeks in duration, prior to screening, according to the criteria defined in the Diagnosis and Statistical Manual of Mental Disorders, Fifth Edition. The diagnosis of MDD will be made by a trained study staff member and supported by the Mini International Neuropsychiatric Interview - Module A.
4. Meet the threshold on the total combined HADS-MADRS score of >/=31 at both screening and day of surgery visits.
5. In sufficiently good health to proceed with planned orthopedic surgery, as ascertained by a standard preoperative clinic evaluation which includes medical history, physical examination (PE), clinical laboratory evaluations, any indicated cardiac testing, and final clearance by the attending anesthesiologists on the day of surgery.
6. Body mass index between 17-40kg/m2.
7. Concurrent psychotherapy will be allowed if the type (e.g., supportive, cognitive behavioral, insight-oriented, et al) and frequency (e.g., weekly or monthly) of the therapy has been stable for at least three months prior to screening and if the type and frequency of the therapy is expected to remain stable during the course of the subject's participation in the study.
8. Concurrent antidepressant therapy (e.g. SSRI or SNRI) and/or hypnotic therapy (e.g. zolpidem, zaleplon, melatonin, or trazodone) will be allowed if the therapy has been stable for at least 4 weeks prior to screening and if it is expected to remain stable during the course of the subject's participation in the study.

<u>Exclusion Criteria:</u>
A potential participant will NOT be eligible for participation in this study if any of the following criteria are met:

1. Female that is pregnant or breastfeeding. These women would not be candidates for elective total joint replacement under any circumstances, and therefore would be screened out from our study population at their routine preoperative evaluation. Women of childbearing potential are routinely screened for pregnancy at their preoperative visit by urine hCG testing if clinically indicated.
2. Total HADS-MADRS score of <31 at either the screening or day of surgery visits.
3. Current diagnosis of a Substance Use Disorder (SUD; Abuse or Dependence, as defined by DSM-V) rated "moderate" or "severe" per criteria of the Mini International Neuropsychiatric Interview – Module J (MINI-J), or Alcohol Use Disorder rated "moderate" or "severe" per MINI-I criteria. These patients are rarely candidates for elective surgery. The following categories of SUD will NOT be excluded: nicotine dependence; alcohol or substance use disorder rated "mild"; alcohol or substance use disorder of any severity in remission, either early (3-12 months) or sustained (>12 months) time frames.
4. History of schizophrenia or schizoaffective disorders, or any history of psychotic symptoms in the current or previous depressive episodes.
5. In the judgment of the investigator, the subject is at significant risk for suicidal behavior during the course of his/her participation in the study.

6. Has dementia, delirium, amnestic, or any other cognitive disorder.
7. Has a clinically significant abnormality on the screening physical examination that would otherwise preclude the patient from having surgery.
8. Participation in any clinical trial with an investigational drug or device within the past month or concurrent to study participation.
9. Lifetime history of surgical procedures involving the brain or meninges, encephalitis, meningitis, degenerative central nervous system disorder (e.g., Alzheimer's or Parkinson's Disease), epilepsy, mental retardation, or any other disease/procedure/accident/intervention associated with significant injury to or malfunction of the central nervous system (CNS), or a history of significant head trauma within the past two years.
10. Presents with any of the following lab abnormalities w/in the past 6 months:
a. Thyroid stimulating hormone (TSH) outside of the normal limits and clinically significant as determined by the investigator. Free thyroxine (T4) levels may be measured if TSH level is high. Subject will be excluded if T4 level is clinically significant.
b. Any other clinically significant abnormal laboratory result at the time of the screening exam.
11. History of hypothyroidism and has been on a stable dosage of thyroid replacement medication for less than six months prior to screening.(Subjects on a stable dosage of thyroid replacement medication for at least six months or more prior to screening are eligible for enrollment.)
12. History of hyperthyroidism which was treated (medically or surgically) less than six months prior to screening.
13. Any current or past history of any physical condition which in the investigator's opinion might put the subject at risk or interfere with study results interpretation.
14. Patients currently maintained on high dose opioids (>90 morphine equivalents per day) prior to surgery.

#### C. Statisical Analysis

Sample Size

Based on published data from other investigators (Zarate et al., 2006) and our own published work (Williams et al., 2018), we estimate a sample size of 15 patients per group, alpha=0.05, power = 80% to detect a 30% change in depression rating from baseline. We will include an additional 5 patients per group to account for participant dropout, for a total of 20 patients per group.

Analysis Plan

1. Statistical analyses will be performed by the investigators using R and GraphPad Prism 9.
2. Baseline characteristics and demographics between groups will be compared using t-tests for continuous and ordinal variables and chi-square tests for categorical variables.
3. Primary outcome: Significance will be determined with a mixed model for repeated measures. MADRS scores obtained on postoperative Day 1, 2, and 3 will be combined and treated as a single timepoint in a mixed model, rather than separately testing each day for significance which can inflate type I error rate. We will use an uncorrected two-sided alpha of 0.05 as a cutoff for statistical significance.
4. Secondary outcomes will be compared using t-tests for continuous and ordinal variables and chi-squared tests for categorical variables. No correction for multiple comparisons needed due to the exploratory nature of these outcomes.

#### D. Study Procedures from Screening to Closeout

Screening

Patients may be informed about the study if they are contacted as part of a perioperative mental health screening service (IRB-54043, approved as not clinical research and not requiring IRB oversight), at which time they may be offered psychiatric care referrals, counseling and information about multiple ongoing clinical trials. Potential subjects may also be introduced to the study through handouts and brochures placed in surgical and pre-anesthesia clinics. Patient identified through Epic as likely to be depressed may also be approached at pre-op appointments and study staff will describe the study and consent if appropriate. Surgery schedulers will ask surgery patients if they are interested in the study and give them a letter or by reading/sending a phone script. Specific procedures for introducing patients to the study and which patient data will be assessed to identify depression are described in detail in section 8g.

Informed consent will be obtained prior to full evaluation of inclusion and exclusion criteria.

Informed consent may be obtained in one of two ways: in-person or over the phone.

In-person consent:

If the patient has an already scheduled preoperative appointment at Stanford, research staff may meet the patient before or after this appointment to obtain informed consent. Alternatively, the patient may choose to make a separate visit to Stanford Medical Center for the consenting process.

Phone consent:

If the patient gave permission for research research staff to contact them about the study (see Section 8g), research staff will call the patient to introduce the study. If the patient agrees to consent, research staff will mail, fax, or email a PDF of the consent form to the patient. Research staff will confirm receipt of the consent form, and then obtain informed consent over the phone. The participant will bring the signed consent form with them during an in-person screening visit, or on the day of surgery or will sign the consent through Redcap. Research staff will verify signatures and sign the researcher signature line at that time.

After providing informed consent, the participant will be offered the option to complete SCREENING in the following ways:

1. During a preoperative visit to Stanford Medicine Outpatient Center.
2. By phone or video interview (using Facetime or Zoom, which is PHI compliant)prior to surgery.
3. In-person visit to Stanford Medical Center.
4. On the day of surgery, prior to surgery at Stanford Hospital; patients will be asked to check in 30-45 minutes early to allow time for screening.

Participants will have the standard preoperative workup and management for surgery, including diagnostic labs/studies and perioperative medication adjustment as needed.

During screening, investigators must document a patient history of Major Depressive Disorder (MDD), single or recurrent. The patient must be currently experiencing a Major Depressive Episode (MDE) of at least eight weeks in duration prior to screening, according to the criteria defined in the Diagnosis and Statistical Manual of Mental Disorders, Fifth Edition. The diagnosis of MDD will be made by a trained study staff member and supported by the Mini International Neuropsychiatric Interview (MINI) - Module A. Patients with a current diagnosis of Alcohol or Substance Use Disorder (Abuse or Dependence, as defined by DSM-V), with the exception of nicotine dependence, at screening or within six months prior to screening will be excluded. The diagnosis of Alcohol or Substance Abuse Disorder will be made by a trained study staff member and supported by the Mini International Neuropsychiatric Interview – Modules I and J.

Concurrent psychotherapy will be allowed if the type (e.g., supportive, cognitive behavioral, insight-oriented, etc.) and frequency (e.g., weekly or monthly) of the therapy has been stable for at least 3 months prior to screening and if the type and frequency of the therapy is expected to remain stable during the course of the subject's participation in the study.

Concurrent antidepressant therapy (e.g. SSRI or SNRI) and/or hypnotic therapy (e.g. zolpidem, zaleplon, melatonin, or trazodone) will be allowed if the therapy has been stable for at least 4 weeks prior to screening and if it is expected to remain stable during the course of the subject's participation in the study.

Once patients agree to participate in the study and have signed the informed consent document, the following screening procedures will be performed:

By Physician or Study Staff:

- Concomitant Medication Review
- Medical and Psychiatric History
- Review of preoperative evaluation data including physical exam, vital signs, any lab work (including a pregnancy test, if applicable) or cardiac testing performed in the course of standard preoperative medical clearance
- Urine toxicology screen test for drugs of abuse
- Mini International Neuropsychiatric Interview (MINI) - Modules A, I & J
- Columbia Suicidality Severity Rating Scale - short screen
- Montgomery-Asberg Depression Rating Scale (MADRS)
- Maudsley Staging Method (MSM)

By the Participant, reviewed by Investigators:

- Hospital Anxiety and Depression Scale (HADS)
- Brief Pain Inventory (BPI)

Participant Compensation:

- $50 after completion of Screening and $50 upon completion of study at day 14. If patients come to Stanford Hospital for the purpose of screening, they will be reimbursed an additional $50 for their time and transportation costs.

<u>Safety Plan for Patients Who Endorse Active Suicidality</u>
A patient may be identified to be at risk for suicidal behavior after consent and during our extended screening process. The Columbia Suicide Severity Rating Scale (CSSRS) – Short Screen is administered face to face, by phone, or by video call (Stanford Zoom) by a study staff member. A patient may be identified as "low", "moderate" or "high" risk. For any patient scoring "moderate" or "high" risk, the study staff will immediately contact Dr. Boris Heifets or Ms. Robin Okada (RN and study coordinator).

If Dr. Heifets or Ms. Okada are notified of a patient at risk for suicidal behavior, Dr. Heifets will contact that patient, verify their response and decide if a wellness check from police is needed or other interventions. That patient will be offered immediate assistance, including calling an ambulance for transport to the Emergency Department, and their primary physician will be informed.

Randomization

Only Investigational Drug Services (IDS) Pharmacy will be aware of drug allocation prior to completion of data collection for the randomized, placebo-controlled study. A computer-generated randomization scheme will be used to randomize participants to receive either ketamine or saline (allocation ratio 1:1, blocks of 4). The study participants, intraoperative anesthesiologists, and study staff conducting postoperative assessments will be blinded to drug allocation until data collection is completed.

Infusions

The study intervention will occur during surgery (Study Day 0). Participants will be randomized to one of two groups: Group A (n=20) will receive a ketamine infusion of 0.5 mg/kg over 40 min during surgery, beginning after anesthetic induction with propofol. Group B (n=20) will receive a saline infusion over 40 min during surgery, beginning after anesthetic induction with propofol. Infusion (instead of bolus) dosing of ketamine was chosen since infusions have demonstrated antidepressant efficacy in outpatient psychiatric populations, as well as theoretically lower risk of adverse events due to lower peak serum concentrations compared to bolus dosing. A patient who is **not** enrolled in the study has a reasonable chance of receiving the same treatments (ketamine or saline) at comparable doses as those tested in the study.

Monitoring

Routine vital signs will be collected before and after surgery by the participant's nurse and reviewed by investigators. Each participant will have continuous physiological monitoring during their anesthetic. A SedLine® Brain Function Monitor will be used to evaluate depth of anesthesia/sedation. If the participant experiences an adverse event temporally related to the infusion where corrective action by the intraoperative anesthesiologist requires unblinding, then the study infusion drug will be revealed to the anesthesiologist.

Data Collection Procedures

The first 5 patients that meet inclusion/exclusion criteria will be entered into the open-label feasibility study; all will receive a ketamine infusion. All other patients enrolled thereafter will be entered into the double-blind, randomized, placebo-controlled study. The day of surgery will be considered Day 0 of the study. Baseline assessments must be performed before the patient receives any psychoactive premedication (e.g. midazolam, fentanyl, gabapentin, oxycodone). Assessments will be performed either in the hospital or, if the patient has been discharged (typically on postoperative Day 2), by phone.

Participants who remain in the hospital for at least 24 hours post-surgery will be asked to give blood samples for immunological analyses. Two blood samples (10mL for each draw) will be collected from each participant.

**Day 0 (Baseline Assessments):**

By Physician or Study Staff:

- Vital Signs
- Concomitant Medication Review
- Adverse Events Review
- Montgomery-Asberg Depression Rating Scale (MADRS)
- Collect breath sample #1

By the Participant, reviewed by Investigators:

- Hospital Anxiety and Depression Scale (HADS)
- Brief Pain Inventory (BPI)

**Day 0 (During Surgery):**
By the Intraoperative Anesthesiologist:
- Ketamine infusion or placebo (saline) infusion
- Continuous physiological monitoring
- SedLine® Brain Function Monitor to evaluate depth of anesthesia/sedation
- Collect breath sample #2, after infusion is complete

**Day 1:**
By Physician or Study Staff:
- Vital Signs
- Concomitant Medication Review
- Adverse Events Review
- Montgomery-Asberg Depression Rating Scale (MADRS)

By the Participant, reviewed by Investigators:
- Hospital Anxiety and Depression Scale (HADS)
- Brief Pain Inventory (BPI)

**Day 2:**
By Physician or Study Staff:
- Vital Signs (inpatient only)
- Concomitant Medication Review
- Adverse Events Review
- Montgomery-Asberg Depression Rating Scale (MADRS)

By the Participant, reviewed by Investigators:
- Hospital Anxiety and Depression Scale (HADS)
- Brief Pain Inventory (BPI)

**Day 3 (inpatient or via phone):**
By Physician or Study Staff:
- Vital Signs (inpatient only)
- Concomitant Medication Review
- Adverse Events Review
- Montgomery-Asberg Depression Rating Scale (MADRS)

By the Participant, reviewed by Investigators:
- Hospital Anxiety and Depression Scale (HADS)
- Brief Pain Inventory (BPI)

**Day 5 (inpatient or via phone):**
By Physician or Study Staff:
- Vital Signs (inpatient only)
- Concomitant Medication Review
- Adverse Events Review
- Montgomery-Asberg Depression Rating Scale (MADRS)

By the Participant, reviewed by Investigators:
- Hospital Anxiety and Depression Scale (HADS)
- Brief Pain Inventory (BPI)

**Day 7 (inpatient or via phone):**

By Physician or Study Staff:
- Vital Signs (inpatient only)
- Concomitant Medication Review
- Adverse Events Review
- Montgomery-Asberg Depression Rating Scale (MADRS)

By the Participant, reviewed by Investigators:
- Hospital Anxiety and Depression Scale (HADS)
- Brief Pain Inventory (BPI)

**Day 14 (inpatient or via phone):**
By Physician or Study Staff:
- Ask participants what treatment they believe they received
- Reveal treatment to participant
- Vital Signs (inpatient only)
- Concomitant Medication Review
- Adverse Events Review
- Montgomery-Asberg Depression Rating Scale (MADRS)

By the Participant, reviewed by Investigators:
- Hospital Anxiety and Depression Scale (HADS)
- Brief Pain Inventory (BPI)

Participant Compensation:
- $50 after completion of all study assessments

Study Procedures Matrix

| | Screen-ing | Day 0 (Baseline) | Day 0 (Intraop) | Day 1 | Day 2 | Day 3 | Day 5 | Day 7 | Day 14 |
|---|---|---|---|---|---|---|---|---|---|
| PHQ-8 | X | | | | | | | | |
| Inclusion/Exclusion Criteria | X | | | | | | | | |
| Demographic data | X | | | | | | | | |
| Consent | X | | | | | | | | |
| Concomitant Medication Review | X | X | | X | X | X | X | X | X |
| Medical and Psychiatric History | X | | | | | | | | |
| Physical Exam | X | | | | | | | | |
| Vital Signs | X | X | | X | X | O | O | O | O |
| Continuous physiological monitoring | | | X | | | | | | |
| Urine toxicology screen test | X | | | | | | | | |
| Labs and studies as warranted by standard preoperative medical clearance (includes pregnancy test) | X | | | | | | | | |
| Mini International Neuropsychiatric Interview (MINI) - A , I & J | X | | | | | | | | |

| | | | | | | | | | |
|---|---|---|---|---|---|---|---|---|---|
| Hospital Anxiety and Depression Scale (HADS) | X | X | | X | X | X | X | X | X |
| Montgomery Asberg Depression Rating Scale (MADRS) | X | X | | X | X | X | X | X | X |
| Columbia Suicide Severity Rating Scale (CSSRS) – Short Screen | X | | | | | | | | |
| Maudsley Staging Method (MSM) | X | | | | | | | | |
| Brief Pain Inventory | X | X | | X | X | X | X | X | X |
| Randomization | X | | | | | | | | |
| Ketamine infusion or Placebo (saline) infusion | | | X | | | | | | |
| SedLine® Brain Function Monitor | | | X | | | | | | |
| Ask participants what treatment they believe they received | | | | | | | | | X |
| Treatment revealed to participant | | | | | | | | | X |
| Participant compensation | X | | | | | | | | X |

X – required
O - inpatient only

#### 9. SIGNIFICANCE

Major Depressive Disorder (MDD) is widely prevalent among patients preparing to have surgery, and is a known risk factor for complications after surgery, including wound infection, myocardial infarction and opioid use disorder. Ketamine has emerged as an effective, rapid-acting antidepressant therapy for patients with MDD, and may be a useful tool to prevent MDD-related morbidity in the perioperative period. Ketamine has been well studied for MDD in outpatient clinics where it is given as an infusion (0.5 mg/kg over 40 minutes) in awake patients.

Ketamine is often used as part of an anesthetic cocktail in sedated or anesthetized patients, but it is unknown whether ketamine has an antidepressant effect in this context. We will determine whether a ketamine infusion, compared to placebo (normal saline infusion), has an antidepressant effect when given during surgical anesthesia. If ketamine is an effective antidepressant in this population under anesthesia, its use could be incorporated into a set of interventions to minimize the perioperative complications associated with MDD.

##### 3. Timeline of Events and Amendments to Protocol

| Event Date | Description of Event/Amendment | |
|---|---|---|
| 3/1/2019 | First submitted study to ClinicalTrials.gov | |
| 3/4/2019 | Original protocol filed with IRB - includes SAP "1.0" and description of open-label feasibility study and RCT | |
| 3/5/2019 | Study posted on ClinicalTrials.gov | |
| 4/12/2019 | Added recruitment letter to patients as a new method of recruitment | REVISION FORM CREATED |
| 4/22/2019 | Changed administrative contact | REVISION FORM CREATED |
| 4/23/2019 | Revised recruitment letter to patients | REVISION FORM CREATED |
| 6/7/2019 | Revised recruitment letter to patients; Added brochure as a new method of recruitment; Added new procedure for approaching potential participants in pre-op appointments; Added method for administering PHQ-8 by filling out a form or over a phone screen | REVISION FORM CREATED |
| 7/24/2019 | First participant consented to the open-label feasibility study | |
| 8/30/2019 | Same as 9/16/19 but the amendment was not yet approved | REVISION FORM CREATED |
| **9/16/2019** | **Inclusion criteria broadened:**<br>**-Type of surgery changed from orthopedic joint to all non-cardiac, non-intracranial surgery**<br>**-Maximum allowable BMI increased from 35 to 40 kg/m2** | |
| 10/8/2019 | Added new personnel to protocol | REVISION FORM CREATED |
| 10/14/2019 | Added another method for administering PHQ-8 by sending through online patient portal | REVISION FORM CREATED |
| **11/6/2019** | **Updated the following eligibility criteria:**<br>**-Added inclusion criterion PHQ8 > 12**<br>**-Changed inclusion criterion for major depressive episode duration from >8 wks to >4 wks**<br>**-Changed allowable concurrent psychotherapy duration from 3 months to 4 weeks** | |
| 11/25/2019 | Removed redundant language in protocol | REVISION FORM CREATED |
| 2/4/2020 | Added new personnel to protocol | REVISION FORM CREATED |
| 2/10/2020 | Last participant consented to the open-label feasibility study | |
| 2/19/2020 | First RCT participant consented but exited study before randomization due to unexpected surgery date change (R001) | |
| 3/16/2020 | Same as 3/26/20 but amendment was not yet approved | REVISION FORM CREATED |
| 3/26/2020 | Clarified phone consent procedures; Modified recruitment brochure; Addition of observation-only arm as an alternative enrollment option (not utilized) | REVISION FORM CREATED |
| 4/2/2020 | Added blood draw to procedures and consent form for a collaborator's study in perioperative immune responses | REVISION FORM CREATED |
| 6/10/2020 | First RCT participant consented and who proceeded to randomization (R002) | |
| 7/6/2020 | Added procedure for telephone consents and signing consent forms via REDCap; Added consent language to allow recording of Zoom assessments for inter-reliability scoring; Added Facetime for assessments; Added thank-you letter for participants; Decreased frequency of blood draws; Changed participant population to allow inclusion of employees | REVISION FORM CREATED |
| 11/20/2020 | Added new personnel to protocol; Removed departed personnel from protoccol; Added gift card confirmation procedures and request for patient address | REVISION FORM CREATED |
| 3/10/2021 | Added new personnel to protocol | REVISION FORM CREATED |
| 4/14/2021 | Added new personnel to protocol | REVISION FORM CREATED |
| 8/9/2021 | Added new personnel to protocol | REVISION FORM CREATED |
| 10/21/2021 | Same as 10/25/22 but amendment was not yet approved | REVISION FORM CREATED |
| 10/22/2021 | Same as 10/25/22 but amendment was not yet approved | |
| 10/25/2021 | Final protocol version filed with IRB containing revised statistical analysis plan | REVISION FORM CREATED |
| 11/4/2021 | Added more descriptive detail to statistical analysis plan (not changing analytic strategy) | REVISION FORM CREATED |
| 12/9/2021 | Added new personnel to protocol | REVISION FORM CREATED |
| 8/11/2022 | Added new personnel to protocol | REVISION FORM CREATED |
| 8/18/2022 | Last RCT participant consented | |
| 8/26/2022 | Last RCT participant randomized and treated | |

4. Original SAP (version 1.0, filed March 4, 2019)

excerpted from protocol version 5, filed March 4, 2019

Stanford Placebo vs Intraoperative Ketamine
Evaluation for Depression
(SPIKED)

**Statistical Analysis Plan Excerpted from Original Protocol**

#### C. Statisical Analysis

Sample Size

Based on published data from other investigators (Zarate *et al*., 2006) and our own published work (Williams *et al*., 2018), we estimate a sample size of 15 patients per group, alpha=0.05, power = 80% to detect a 30% change in depression rating from baseline. We will include an additional 5 patients per group to account for participant dropout, for a total of 20 patients per group.

Analysis Plan

1. Statistical analyses will be performed by the investigators using GraphPad Prism 8.
2. Baseline characteristics and demographics between groups will be compared using t-tests for continuous and ordinal variables and chi-square tests for categorical variables.
3. Primary outcome: superiority of ketamine to saline infusion will be demonstrated by a statistically significant greater decrease in the composite score of the Hospital Anxiety and Depression Scale (HADS) and Montgomery and Asberg Depression Rating Scale (MADRS) when comparing baseline, pre-infusion scores to postoperative scores, using a t-test ($p<0.05$, 2-sided, adjusting for multiple comparisons).
4. Secondary outcomes between groups will be compared using t-tests for continuous and ordinal variables and chi-squared tests for categorical variables.

5. Final SAP (version 2.0, filed November 4, 2021)
created from protocol version 7, filed October 22, 2021

Stanford Placebo vs Intraoperative Ketamine
Evaluation for Depression
(SPIKED)

### Statistical Analysis Plan (SAP)

#### Section 1: Administrative Information

##### 1. Title and trial registration

**1a. Title**
Intraoperative Ketamine Versus Saline in Depressed Patients Undergoing Anesthesia for Non-Cardiac Surgery

**1b. Trial registration number**
ClinicalTrials.gov Identifier NCT# 03861988

**2. SAP version:** 2.0

**3. Protocol version:** Stanford eProtocol ID 49114, last revision submitted 8/9/21

##### 4. SAP revisions

**4a. SAP revision history**
10/21/2019 – version 1.0 (originally included in the study protocol)
10/28/2021 – version 2.0 (created a new, separate SAP document)

**4b. Justification for each SAP revision**
**Version 1.0 to 2.0:**

1) In our original protocol, recruitment was limited to patients with symptomatic depression presenting for hip and knee arthroplasty at Stanford. Patients undergoing this type of surgery have a predictable postoperative course, which guided our primary endpoint selection. We encountered significant difficulties recruiting patients during the COVID-19 pandemic. Therefore, we expanded eligibility criteria by including patients presenting for a wider variety of surgical procedures, which are described in our updated protocol. To account for the likely increased variability of postoperative recovery in this broader population, we updated our primary endpoint to include data from the first 3 days after surgery, instead of only a single day after surgery.
2) In our original protocol, the primary outcome measure was defined as the sum of scores from a clinician-administered scale and a self-report scale. During the recruitment phase of our trial, and prior to unblinding, we presented our analysis plan to a panel of statisticians in the Department of Biomedical Data Sciences at Stanford University. The panel recommended that we could improve the interpretability and reduce the variance in our primary outcome by only using one validated measure (rather than a composite score) as our primary outcome measure. Based on this recommendation, we updated our primary outcome measure to only include the clinician-administered scale (the Montgomery-Asberg Depression Rating Scale).

##### 4c. Timing of SAP revisions

This revision (1.0 to 2.0) was made prior to data unblinding and prior to completion of data collection (75% of target enrollment).

#### 5. Roles of SAP contributors

Theresa R. Lii, MD, Stanford University, Primary SAP author
Boris D. Heifets, MD, PhD, Stanford University, Chief investigator/clinical lead

#### 6. Signatures

**6a. Primary SAP author:** 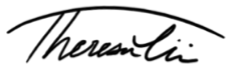

**6b. Chief investigator/clinical lead:** 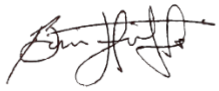

#### Section 2: Introduction

##### 7. Background and rationale

Major depressive disorder (MDD) is widely prevalent among patients preparing to have surgery, and is a known risk factor for complications after surgery, including wound infection, myocardial infarction and opioid use disorder. Ketamine has emerged as an effective, rapid-acting antidepressant therapy for patients with MDD, and may be a useful tool to prevent MDD-related morbidity in the perioperative period. Ketamine has been well studied for MDD in outpatient clinics where it is given as an infusion (0.5 mg/kg over 40 minutes) in awake patients. Ketamine is often used as part of an anesthetic cocktail in sedated or anesthetized patients, but it is unknown whether ketamine has an antidepressant effect in this context. We will determine whether a ketamine infusion, compared to placebo (normal saline infusion), has an antidepressant effect when given during surgical anesthesia.

##### 8. Objectives

We hope to discover whether ketamine has antidepressant efficacy in major depressive disorder (MDD) patients when given as an anesthetic adjunct. If ketamine is an effective antidepressant in this population under anesthesia, its use could be incorporated into a set of interventions to minimize the perioperative complications associated with MDD. While it is more efficient to deliver ketamine therapy intraoperatively, if this study finds that ketamine is ineffective in this setting, that result establishes a rationale to test treatment prior to the surgical encounter. We will also have gained important information on ketamine's antidepressant mechanism (e.g. it is blocked by other anesthetics, or it requires that the patient be conscious).

#### Section 3: Study Methods

##### 9. Trial design

This is a single-site phase IV randomized clinical trial utilizing a quadruple-masked, placebo-controlled, parallel arm design. Participants will be randomly allocated in a 1:1 ratio to one of two groups: Group A (n=20) will receive a ketamine infusion of 0.5 mg/kg over 40 min during surgery. Group B (n=20) will receive a saline infusion over 40 min during surgery. In both groups, the study drug will be given after anesthetic induction. The participants, care providers, investigators, and outcomes assessors will be masked.

##### 10. Randomization

Participants will be sequentially randomized as they enter the study. We will employ block randomization with 5 blocks of 8 patients each.

##### 11. Sample size

We will enroll a total of 40 participants. Our sample size calculation was derived from an *a priori* power analysis for the primary outcome measure with a 1:1 allocation ratio. In a randomized controlled trial of ketamine vs active placebo (midazolam) performed by

Phillips et al. (2019), participants had a mean decrease of 10.9 points (SD=8.9) in MADRS total score relative to pre-infusion scores compared with a mean decrease of 2.8 points (SD=3.6) with midazolam. Using these results, we computed an estimated total sample size of 38 participants at a two-sided alpha level of 0.05 and 80% power. An additional 2 participants were added to account for potential attrition, for a total of 40 participants. Given the increased statistical efficiency of mixed models for repeated measures (MMRM) compared to parametric pre-post tests, we assumed that 40 participants should be sufficient to power our study for MMRM analysis.

**12. Framework**
We plan to evaluate the antidepressant superiority of ketamine to placebo by assessing the postoperative MADRS scores from postoperative Day 1 through Day 3. Additional outcomes collected at postoperative Day 5, 7, and 14 will be used for statistical model building and exploratory analyses.

**13. Interim data monitoring**

**13a. Statistical interim analyses planned and time points**
No statistical interim analyses planned.

**13b. Any planned adjustment of significance level due to interim analysis**
N/A.

**13c. Details of guidance for stopping the trial early**
N/A.

**14. Timing of final analysis**
Once all data have been collected at all planned time points from the final (40th) participant, all investigators will be unmasked and all outcomes will be analyzed together in the final analysis.

**15. Timing of outcome assessments**
All patients will have at least one baseline psychiatric assessment prior to Day 0. Repeat baseline assessment by questionnaire will occur on the morning of surgery prior to any preoperative medication being given. The study assessments will be performed at postoperative Day 1, Day 2, Day 3, Day 5, Day 7 and Day 14. Assessments will be performed either in hospital or by phone or video.

#### Section 4: Statistical Principles

**16. Level of statistical significance**
We will use an uncorrected two-sided alpha of 0.05 as a cutoff for statistical significance.

**17. Adjustment for multiplicity and controlling type I error**

The primary outcome (MADRS score) that is measured on postoperative Day 1, 2, and 3 will be combined and treated as a single timepoint in the mixed model, rather than separately testing each day for significance which can inflate type I error rate. Postoperative Days 1, 2, and 3 were chosen due to previously published data showing that the antidepressant effect of ketamine persists up to 3 days after administration.

**18. Confidence intervals to be reported**
We will report 95% confidence intervals for the primary outcome.

**19. Adherence and protocol deviations**

**19a. Adherence to protocol and how this is assessed**
Adherence to protocol is defined as: 1) participant meets eligibility criteria prior to randomization; 2) all planned assessments are completed; and 3) the patient receives the complete dose of the blinded study drug intravenously over a course of 40 minutes, initiated after induction of anesthesia and completed before extubation or transfer to the post-anesthetic recovery area.

**19b. How adherence to protocol will be presented**
Adherence to protocol will be presented as a proportion.

**19c. Protocol deviations**
Emergency protocol deviations are defined as those occurring in an emergency situation where departure from the protocol (such as unblinding the study drug) is required immediately to protect the life or physical well-being of the participant.

Non-emergent deviations are defined as those that do not occur in an emergency situation. Examples potentially encountered in our study include, but are not limited to: 1) participant is randomized to a treatment intervention but does not meet eligibility criteria, 2) the anesthesiologist administered the study drug incorrectly or incompletely, 3) the participant inadvertently received the intervention meant for the other trial arm, 4) the participant was not available for assessment of the planned outcome either because of loss to follow-up or for another reason, 5) study staff were not available for assessment of the planned outcome either because of a personal emergency or another reason.

**19d. Which protocol deviations will be summarized**
All emergency and non-emergent protocol deviations will be summarized.

**20. Analysis populations**
As recommended by CONSORT guidelines for parallel group randomized controlled trials, intention-to-treat and per-protocol analyses (if needed) will be performed and reported for all planned outcomes to allow readers to interpret the effects of the intervention.

**<u>Section 5: Trial Population</u>**

#### 21. Screening data

Screening data to be collected and reported in CONSORT flow diagram include:

Patient Health Questionnaire score
HADS-MADRS composite score
English-speaking status
Age
Body mass index
History of significant CNS disease or brain surgery
History of moderate to severe substance use disorder (except nicotine)
History of schizophrenia, schizoaffective disorder, or psychosis
Chronic high dose opioid use (>90 morphine milligram equivalents per day)
High suicide risk
Other reasons

#### 22. Eligibility

##### 22a. Inclusion criteria

A subject will be eligible for inclusion only if all of the following criteria are met:

1. Male or female, 18 to 80 years of age, inclusive, at screen.

2. Able to read, understand, and provide written, dated informed consent prior to screening. Participants will be deemed likely to comply with study protocol and communicate with study personnel about adverse events and other clinically important information.

3. PHQ-8 score >= 12.

4. Diagnosed with Major Depressive Disorder (MDD), single or recurrent, and currently experiencing a Major Depressive Episode (MDE) of at least 4 weeks in duration, prior to screening, according to the criteria defined in the Diagnosis and Statistical Manual of Mental Disorders, Fifth Edition. The diagnosis of MDD will be made by a trained study staff member and supported by the Mini International Neuropsychiatric Interview - Module A.

5. Meet the threshold on the total combined HADS-MADRS score of >/=31 at both screening and day of surgery visits.

6. In sufficiently good health to proceed with planned surgery, as ascertained by a standard preoperative clinic evaluation which includes medical history, physical examination (PE), clinical laboratory evaluations, any indicated cardiac testing, and final clearance by the attending anesthesiologists on the day of surgery.

7. Body mass index between 17-40kg/m2.

8. Concurrent psychotherapy will be allowed if the type (e.g., supportive, cognitive behavioral, insight-oriented, et al) and frequency (e.g., weekly or monthly) of the therapy has been stable for at least 4 weeks prior to screening and if the type and frequency of the therapy is expected to remain stable during the course of the subject's participation in the study.

9. Concurrent antidepressant therapy (e.g. SSRI or SNRI) and/or hypnotic therapy (e.g. zolpidem, zaleplon, melatonin, or trazodone) will be allowed if the therapy has been stable for at least 4 weeks prior to screening and if it is expected to remain stable during the course of the subject's participation in the study.

**22b. Exclusion criteria**
A potential participant will NOT be eligible for participation in this study if any of the following criteria are met:

1. Female that is pregnant or breastfeeding. These women would not be candidates for elective surgery under any circumstances, and therefore would be screened out from our study population at their routine preoperative evaluation. Women of childbearing potential are routinely screened for pregnancy at their preoperative visit by urine hCG testing if clinically indicated. We will not be including pregnant women from other surgical populations as well.

2. Total HADS-MADRS score of <31 at either the screening or day of surgery visits.

3. Current diagnosis of a Substance Use Disorder (SUD; Abuse or Dependence, as defined by DSM-V) rated "moderate" or "severe" per criteria of the Mini International Neuropsychiatric Interview – Module J (MINI-J), or Alcohol Use Disorder rated "moderate" or "severe" per MINI-I criteria. These patients are rarely candidates for elective surgery. The following categories of SUD will NOT be excluded: nicotine dependence; alcohol or substance use disorder rated "mild"; alcohol or substance use disorder of any severity in remission, either early (3-12 months) or sustained (>12 months) time frames.

4. History of schizophrenia or schizoaffective disorders, or any history of psychotic symptoms in the current or previous depressive episodes.

5. In the judgment of the investigator, the subject is at significant risk for suicidal behavior during the course of his/her participation in the study.

6. Has dementia, delirium, amnestic, or any other cognitive disorder.

7. Has a clinically significant abnormality on the screening physical examination that would otherwise preclude the patient from having surgery.

8. Participation in any clinical trial with an investigational drug or device that conflicts with this trial, within the past month or concurrent to study participation.

9. Lifetime history of surgical procedures involving the brain or meninges, encephalitis, meningitis, degenerative central nervous system disorder (e.g., Alzheimer's or Parkinson's Disease), epilepsy, mental retardation, or any other disease/procedure/accident/intervention associated with significant injury to or malfunction of the central nervous system (CNS), or a history of significant head trauma within the past two years.

10. Presents with any of the following lab abnormalities w/in the past 6 months:
    a. Thyroid stimulating hormone (TSH) outside of the normal limits and clinically significant as determined by the investigator. Free thyroxine (T4) levels may be measured if TSH level is high. Subject will be excluded if T4 level is clinically significant.
    b. Any other clinically significant abnormal laboratory result at the time of the screening exam.

11. History of hypothyroidism and has been on a stable dosage of thyroid replacement medication for less than six months prior to screening. (Subjects on a stable dosage of thyroid replacement medication for at least six months or more prior to screening are eligible for enrollment.)

12. History of hyperthyroidism which was treated (medically or surgically) less than six months prior to screening.

13. Any current or past history of any physical condition which in the investigator's opinion might put the subject at risk or interfere with study results interpretation.

14. Patients currently maintained on high dose opioids (>90 morphine equivalents per day) prior to surgery

**23. Recruitment**
Information to be collected and reported in CONSORT flow diagram include:

Patients with a history of depression screened
Excluded due to PHQ score < 12
Excluded due to other exclusion criteria
Declined to participate
Lost to follow-up during screening
Excluded after consent
Withdrew after consent prior to intervention
Participants consented

#### 24. Withdrawal and follow-up

##### 24a. The following withdrawal/lost to follow-up data will be recorded:

- Number of participants who withdraw or are lost to follow-up
- Timing of withdrawal or lost to follow-up
- Reasons of withdrawal or lost to follow-up

##### 24b. The above withdrawal/lost to follow-up data will be presented as:

- Percentages (%) of participants who withdraw or are lost to follow-up
- Kaplan-Meier curves of withdrawal or lost to follow-up (if any)
- Reasons of withdrawal or lost to follow-up summarized in the Results section

#### 25. Baseline patient characteristics

##### 25a. List of baseline characteristics to be reported

1) Sex (n, %)
2) Age (years, mean and SD)
3) Body mass index (mean, SD)
4) Hispanic/Latino ethnicity (n, %)
5) Race
    a. White (n, %)
    b. Black/African American (n, %)
    c. Asian (n, %)
    d. Native Hawaiin/Pacific Islander (n, %)
    e. Other (n, %)
    f. Unknown (n, %)
6) Married (n, %)
7) Disabled (n, %)
8) Current smoker (n, %)
9) PHQ score at screening (points, mean and SD)
10) HADS at screening (points, mean and SD)
11) MADRS at screening (points, mean and SD)
12) HADS-MADRS composite score at screening (points, mean and SD)
13) Age at first MDD onset (years, mean and SD)
14) Duration of current episode (months, mean and SD)
15) Recurrent major depression (n, %)
16) Previous clinical ketamine exposure (n, %)
17) Previous recreational ketamine exposure (n, %)
18) Charlson Comorbidity Index (points, mean and SD)
19) Maudsley resistance stage (points, mean and SD)
20) Numerical pain score (points, mean and SD)
21) Pain interference from BPI (points, mean and SD)

##### 25b. Preoperative psychoactive medications

1) Opioids (n, %)

2) MME/day, if on opioids (mean, SD)
3) Anticonvulsants (n, %)
4) Benzodiazepines (n, %)
5) Muscle relaxants (n, %)
6) Antidepressants (n, %)
7) Antipsychotics (n, %)
8) Lithium (n, %)
9) Prescription sleep medications (n, %)
10) Opioid antagonists (n, %)
11) Stimulants (n, %)

**25c. Surgical characteristics**

1) Type of surgery
    a. ENT/OMF(n, %)
    b. Thoracic (n, %)
    c. Breast (n, %)
    d. Plastic non-breast (n, %)
    e. Spine (n, %)
    f. Major joint (n, %)
    g. Other orthopedic/limb (n, %)
    h. Intraabdominal general surgery (n, %)
    i. Urologic and gynecologic (n, %)
    j. Vascular (n, %)
    k. Transplant (n, %)
2) Type of anesthesia
    a. General anesthesia (n, %)
    b. Monitored anesthesia care (n, %)
3) Intraoperative nitrous oxide (n, %)
4) Intraoperative propofol infusion (n, %)
5) Intraoperative opioid infusion (n, %)
6) Length of surgery (hours, mean and SD)
7) Peripheral or neuraxial catheter placed (n, %)

**Section 6: Analysis**

**26. Outcome definitions**

**26a. Primary outcome**

Postoperative MADRS scores from postoperative Days 1 through 3.

**26b. Main secondary outcomes**

1) Proportion of participants with clinical response (defined as a ≥50% reduction in MADRS score from baseline).
2) Proportion of participants with remission (defined as a MADRS score of ≤12 on day 14).

##### 26c. Other exploratory secondary outcomes

1) MADRS scores at postoperative Days 5, 7, and 14
2) Hospital Anxiety and Depression Scale
3) Cumulative inpatient opioid use
4) Hospital length of stay
5) Average inpatient opioid use per day
6) Opioid use at postop day 7
7) Opioid use at postop day 14
8) Postoperative numeric pain scores
9) Postoperative pain interference scores

#### 27. Analysis methods

##### 27a. Analysis of the primary outcome

A mixed model for repeated measures (MMRM) will be used to analyze the difference in postoperative MADRS score which is our primary outcome. The model will include treatment and time as fixed effects, with a treatment*time interaction term. The data will be modeled with random intercepts and random slopes to account for variation in baseline scores and postoperative trajectories. MADRS scores measured on postoperative Day 1, 2, and 3 will be combined and treated as a single timepoint in the mixed model, rather than separately testing each day for significance which can inflate type I error rate. Postoperative Days 1, 2, and 3 were chosen due to previously published data showing that the antidepressant effect of ketamine persists up to 3 days after administration.

##### 27b. Sensitivity analyses

Sensitivity analyses will be conducted for the following scenarios:

1) Missing data: See section 28
2) Intention-to-treat vs per-protocol analysis (if needed)
3) MMRM analysis using HADS (self-report) as a confirmatory measure
4) MMRM analysis using the previously proposed primary outcome measure (a composite score defined as the sum of scores from MADRS and HADS)

##### 27c. Subgroup analyses

We will conduct several pre-specified subgroup analyses. While the trial will not specifically be powered for these subgroups, we hope to gain further insight into sub-populations that may particularly benefit from the intervention. Example subgroup analyses to be considered:

1) Preoperative use of antidepressants
2) Preoperative use of opioids
3) Intraoperative exposure to nitrous oxide
4) Intraoperative exposure to opioid infusions
5) Postoperative use of neuraxial or peripheral nerve catheters

**28. Missing data**
For the primary outcome, it will be assumed that the missing data are missing at random. Sensitivity analysis will be performed to evaluate the robustness of the MMRM to the missing-at-random assumption.

**29. Adverse events**
Adverse events will be recorded in the Stanford REDCap database using the Adverse Event (AE) Form and Serious Adverse Event (SAE) Form. Refer to document GUI-P13 (https://researchcompliance.stanford.edu/panels/hs/policies/guidances) for definitions of AEs and SAEs. All AEs and SAEs will be summarized.

**30. Statistical software**
Statistical analyses will be conducted using R statistical software. Data figures for visualization will be generated using either R or Graphpad Prism.

**6. Timeline of Events and Amendments to Statistical Analysis Plan (SAP)**

| Event Date | Description of Event/Amendment |
|---|---|
| 3/4/2019 | Original protocol filed with IRB - includes SAP "1.0" and description of open-label feasibility study and RCT |
| **11/6/2019** | **Updated primary endpoint to postoperative day 1 as primary endpoint based on open-label feasibility study data. Protocol updated and submitted to IRB.** |
| **10/22/2021** | **Final protocol version filed with IRB after consultation with Stanford Data Studio statisticians:<br>-Primary outcome changed from MADRS+HADS composite score to MADRS score to bring into conformity with other ketamine RCTs<br>-Primary endpoint changed from postoperative day 1 to postoperative days 1, 2 and 3 analyzed with mixed models for repeated measures. Primare endpoint timeframe changed due to high variance observed at day 1. Change to mixed model as this analysis is more sensitive to detect a between-group difference versus parametric test.** |
| 10/28/2021 | Created a final SAP document separated from study protocol (version "2.0") |
| 11/4/2021 | Filed the final version of SAP "2.0" to IRB |
